# Late-Pregnancy Immune and Kynurenine Signatures Predict Postpartum Depression and Brain Morphology

**DOI:** 10.64898/2026.10.02.26364495

**Authors:** Yasmin A. Harrington, Ha Kyung Shin, Chiara Milano, Massimo C. Mauri, Alessandra Marano, Eleonora Maggioni, Letizia Squarcina, Emma Tassi, Paolo Enrico, Nicola Persico, Giorgio Conte, Elisa M.T Melloni, Elena Agnoletto, Sara Spadini, Cristina Lorenzi, Sofia Nasini, Stefano Dall’Acqua, Stefano Comai, Sara Poletti, Massimo Origoni, Paolo Brambilla, Francesco Benedetti, Milano PPDINFLAME Study Group

**Affiliations:** Psychiatry & Clinical Psychobiology, IRCCS Scientific Institute Ospedale San Raffaele, Milan, Italy; Vita-Salute San Raffaele University, Milan, Italy; Department of Neurosciences and Mental Health, Fondazione IRCCS Ca’ Granda, Ospedale Maggiore Policlinico, Milan, Italy; Department of Pathophysiology and Transplantation, University of Milan, Milan, Italy; Department of Electronics Information and Bioengineering, Politecnico di Milano, Milano, Italy; Fetal Medicine and Surgery Service, Fondazione IRCCS Ca’ Granda Ospedale Maggiore Policlinico, Milan, Italy; Department of Neuroradiology, Fondazione IRCCS Ca’ Granda, Ospedale Maggiore Policlinico, Milan, Italy; Department of Pharmaceutical and Pharmacological Sciences, University of Padua, Padua, Italy; Department of Psychiatry, McGill University, Montreal, QC, Canada; Department of Gynecology & Obstetrics, IRCCS Ospedale San Raffaele, Milano, Italy

**Author notes:** **Corresponding author**: Yasmin A. Harrington.

**Keywords:** Postpartum depression, pregnancy, inflammation, prediction, neuroimaging, biomarkers

## Abstract

**Background:** Pregnancy involves profound immune adaptations that may contribute to the development of postpartum depression (PPD); however, the relevant biomarkers and critical time window remain unclear. Therefore, we investigated whether third-trimester and postpartum cytokines and kynurenine pathway (KP) metabolites predict PPD and whether these biological signatures are related to postpartum brain morphology.

**Methods:** Circulating cytokines and KP metabolites were assessed in 140 pregnant women during the third trimester and postpartum. Partial least squares (PLS) regression models were used to identify multivariate biological profiles predictive of PPD, followed by classification and regression tree (CART) analyses to evaluate predictive performance. In a neuroimaging subsample (n=61), voxel-based morphometry examined associations among PPD status, significant PLS-derived components, selected individual biomarkers, and postpartum grey matter volume.

**Results:** A third-trimester cytokine signature including 21 cytokines significantly predicted PPD with high specificity. A third-trimester KP profile, driven primarily by the kynurenic acid/quinolinic acid ratio, also predicted PPD, and the combination of cytokine and KP components improved overall predictive performance to 93.3%. Postpartum biomarkers showed substantially weaker associations with PPD. The cytokine component was differentially associated with postpartum grey matter volume according to PPD status, with opposite associations observed in occipital regions between women who did and did not develop PPD. KP measures also showed distinct associations within sensorimotor regions based on PPD status

**Conclusions:** Late pregnancy immune-inflammatory alterations are associated with subsequent PPD and postpartum brain morphology, supporting the potential utility of pregnancy-derived biological signatures for identifying women at increased risk for PPD.

## 1. INTRODUCTION

Postpartum depression (PPD) is a common complication after childbirth, with systematic reviews estimating a global prevalence higher than 20% (1), and with 6.2% of women developing a full-blown major depressive disorder (MDD) during the peripartum period (2). Symptoms largely resemble those of MDD, yet PPD is distinct in its timing of onset. One third of PPD cases develop during pregnancy and in roughly 25-50% of all cases, symptoms can persist longer than 6 months, requiring antidepressant treatment (3). Moreover, suicide is the leading cause of maternal death in the first year after delivery (4). PPD is a tremendous burden for the mother and can negatively influence parenting behavior (5) and impact cognitive and emotional development in offspring (6–8). Therefore, there is an urgent need to identify biomarkers and predictors of illness onset, to identify women at high risk of developing early postnatal depression and support targeted preventive interventions.

Evidence increasingly implicates immune dysregulation including systemic monocyte-macrophage activation, low-grade inflammation through higher levels of circulating pro-inflammatory cytokines and kynurenine pathway (KP) metabolites, and premature T cell defects in PPD (9). During the peripartum period, innate and adaptive immunity undergoes drastic changes to accommodate the growing fetus, with depressive symptoms being the most prevalent during the third trimester and postpartum, when the mother’s immune system is in an inflammatory state (10, 11). The majority of studies have associated higher levels of immune-inflammatory signaling compounds during pregnancy with antepartum depression, and higher postpartum levels with PPD (9, 10, 12–20). Moreover, lower levels of anti-inflammatory factors during pregnancy have also been associated with subsequent PPD (21), although inflammatory markers have shown heterogeneous and occasionally opposing associations across studies (22, 23). While systematic reviews support an overall association between inflammatory markers and peripartum depression (12), negative findings remain frequent. Results appear more consistent for antepartum depression than for PPD especially when utilizing postpartum cytokines (24). Few studies have evaluated the predictive capacity of pregnancy immune profiles and individual biomarkers have generally shown limited predictive performance for PPD (20).

These inconsistencies may be due to different study methods, but also to the selection of cytokines as predictors that are most associated with pregnancy-unrelated MDD (10, 12, 24, 25). It can be surmised that the relationship between cytokines and mood might be different in pregnancy, postpartum, and in pregnancy-unrelated MDD; with unbalanced immune changes during pregnancy fostering both, psychiatric and medical conditions (9, 26). In addition, the KP provides a biologically relevant link between immune activation and brain function as inflammatory signaling can increase tryptophan catabolism and alter the balance among neuroactive kynurenine metabolites. In particular, kynurenic acid (KYNA) and quinolinic acid (QUIN) exert divergent effects on glutamatergic and neuroinflammatory signaling (27). Examining cytokines and KP metabolites jointly may therefore capture coordinated biological alterations that cannot be adequately represented by single-biomarker approaches.

Additionally, grey matter volumes (GMV) undergo change in the perinatal period. Normal pregnancy involves widespread reductions in GMV that normalize or increase postpartum (28–31), but can also persist over several years (32). Conversely, researchers have found increases in GMV in areas related to maternal behavior including the amygdala, hypothalamus, and substantia nigra (33, 34). Neuroimaging studies of PPD remain limited and heterogenous yet, some studies have reported larger GMV in women with PPD compared to healthy controls (HC) (35–37), opposite of what is generally found in non-perinatal depression (38). Cytokine levels have been negatively associated with GMV in non-perinatal populations, suggesting a possible relationship between immune signaling and brain structure (39), but no study has directly investigated the relationship between perinatal inflammatory markers and postpartum GMV.

Therefore, this study aimed to investigate whether multivariate profiles derived from a large panel of circulating inflammatory markers and KP metabolites were associated with subsequent PPD status, and whether these biological profiles showed different relationships with postpartum GMV according to PPD status in a homogeneous sample of women without a previous history of major psychiatric disorders, recruited in an outpatient real-world setting.

## 2. METHODS AND MATERIALS

### 2.1 Participants

We studied 140 pregnant women enrolled between 2022 and 2025 at Department of Obstetrics and Gynecology of IRCCS San Raffaele Hospital and Fondazione IRCCS Ca’ Granda Ospedale Maggiore Policlinico in Milan (Italian Ministry of Health, PPDINFLAME project RF-2019-12371066; ClinicalTrials.gov ID NCT05784116). PPD was assessed using the Edinburgh Postnatal Depression Scale (EPDS), administered during the third trimester of pregnancy and 6 weeks postpartum. PPD status was determined based on a cutoff score of 11 and above on the EPDS, which had the best AUC at ROC analysis in the Italian population (40, 41). Women with EPDS≥11 were categorized as having probable PPD, mentioned in text as PPD for simplicity, and women with EPDS≤10 were categorized as HC. Individuals were excluded if under 18 or over 50 years old, had a history of bipolar disorder, schizophrenia, schizoaffective disorder, psychosis; anorexia or bulimia nervosa; took any of the following medications: antipsychotics, anticonvulsants, mood stabilizers, stimulants; had alcohol and/or substance dependence or abuse in the last 6 months, had any major medical and neurological disorders or medical conditions affecting the immune system, or ongoing drug treatment known to affect the immune system (i.e. anti-inflammatory agents, corticosteroids, immunosuppressants). Written informed consent was obtained from participants after a full explanation of the study, as approved by the local Ethics Committees.

### 2.2 Analysis of peripheral markers of immune-inflammatory activation and KP

Blood was collected by venipuncture into EDTA-containing Vacutainer tubes in the morning after an overnight fast, then centrifuged at 2000 × g for 15 min at 4°C, and plasma was aliquoted and stored at −80°C to investigate levels of peripheral inflammatory markers. Luminex Bio-plex Pro Human Cytokine 48-Plex immunoassays were run to determine plasma concentrations of immune markers using the bead-based Luminex system with xMAP technology (Luminex 200 system, Merck Millipore). Analyses were performed using concentrations of observed markers (pg/ml) calculated using Belysa Immunoassay software (version 1.2).

The 48 inflammatory markers included were Interleukin 1α IL-1α; IL-1β; Interleukin-1 receptor antagonist IL-1ra; Interleukin-2 receptor α chain IL-2Rα (CD25); IL-2; IL-3; IL-4; IL-5; IL-6; IL-7; IL-8; IL-9; IL-10; IL-12 (p40); IL-12 (p70); IL-13; IL-15; IL-16; IL 17A; IL-18; Fibroblast growth factor FGF basic; Eotaxin; Granulocyte colony-stimulating factor G-CSF; Granulocyte-macrophage colony-stimulating factor GM-CSF; Interferon-γ IFN-γ; IFN-α2; Growth-regulated oncogene α GRO-α (CXCL1); Hepatocyte growth factor HGF; Leukemia inhibitory factor LIF; Monocyte chemoattractant protein 1 MCP-1 (MCAF; CCL2); Monocyte chemoattractant protein 3 MCP-3; Interferon gamma-induced protein 10 IP-10 (CXCL10); Monokine Induced by Gamma interferon MIG; Nerve growth factor β-NGF; Stem cell factor SCF; Stem Cell Growth Factor-β SCGF-β; Stromal cell-derived factor 1 SDF-1α; Macrophage Inflammatory Protein-1α MIP-1α (CCL3); MIP-1β (CCL4); Platelet-Derived Growth Factor-BB PDGF-BB; Regulated on Activation Normal T-cell Expressed and Secreted RANTES; Vascular Endothelial Growth Factor VEGF; Cutaneous T-cell-attracting chemokine CTACK; Macrophage migration inhibitory factor MIF; TNF-related apoptosis-inducing ligand TRAIL; Macrophage colony-stimulating factor M-CSF; Tumor Necrosis Factor TNF-α; and TNF-β (Lymphotoxin α).

KP metabolites were quantified using previously validated analytical methods (42, 43). Plasma concentrations of tryptophan (TRP), kynurenine (KYN), 3-hydroxykynurenine (3-HK), kynurenic acid (KYNA), and quinolinic acid (QUIN) were determined using HPLC with UV and fluorometric detection and LC-MS/MS. Metabolite ratios were calculated as indirect indices of KP enzyme activity, including KYN/TRY (indoleamine 2,3-dioxygenase and tryptophan 2,3-dioxygenase activity), KYNA/KYN (kynurenine aminotransferase II activity), and 3-HK/KYN (kynurenine-3-monooxygenase activity). The KYNA/QUIN ratio was also calculated as an index of relative balance between neuroprotective and neurotoxic kynurenine metabolism.

### 2.3 Multivariate Statistical analysis

To select factors of interest in predicting PPD, we used partial least squares regression (PLS), a machine learning technique that models the relationships between sets of observed variables with latent variables, to define a linear regression model by projecting the variables into a new space. PLS creates composite variables, commonly referred to as “scores” or latent components, from the predictor data matrix (X). PLS uses information from the response (Y) to derive latent components that maximize the covariance between predictors and response (44). Separate analyses were performed for two predictor blocks due to differences in data availability: (1) cytokines, chemokines, and growth factors, and (2) KP metabolites. In both models, the predictor matrix (X) consisted of the analytes included in the performer biological block, whereas the response (Y) was postpartum PPD status coded as [0 = EPDS<11 and 1 = EPDS≥11]. The optimal number of latent components (A) was selected through K-fold cross validation with 7 folds.

Model performance was summarized using R^2^X (an A-dimensional vector, to record the explained variance of the predictors by each PLS component), R^2^Y (an A-dimensional vector, to record the explained variance of the response variables by each PLS component), and Q^2^ (predicted variation, to measure R^2^Y applied to a test set with cross-validation procedure). The contribution of each predictor variable was described using PLS weights (w) and the variable importance in projection (VIP) scores. Variables with VIP>1 were considered to provide an above-average contribution to the multivariate model (45–48). Participants with valid measurements for fewer than 80% of analytes were excluded, as were analytes available in fewer than 80% of participants. Recruitment center, age, number of previous pregnancies, and gestational week (for third trimester concentrations) or days from delivery to blood sampling (for postpartum concentrations) were also entered into the predictor matrix.

Then, for explanatory purposes, the analytes with VIP>1 at the PLS were entered separately as independent variables into a GLZM analysis of homogeneity of slopes to predict PPD (49). Parameter estimates were obtained with iterative re-weighted least squares maximum likelihood procedures. The significance of the effects was calculated with likelihood ratio (LR) statistics (50, 51). This PLS+GLZM has consistently been able to reveal complex genetic and immune-brain interactions in mood disorders, including PPD (52–55).

We then evaluated whether the individual scores of the predictive variable defined by the PLS algorithm were able to provide a 50% higher performance when used in a binary classifier to predict the presence/absence of PPD, using a decision tree based on the classification and regression tree (CART) algorithm (56), a non-parametric supervised learning method that constructs binary trees by recursively partitioning data and using the Gini Impurity index to identify the optimal split that maximizes homogeneity within resulting segments. The resulting classification matrix was then evaluated for sensitivity, specificity, predictive values, and accuracy, at an estimated prevalence of PPD=20%.

All analyses were performed with StatSoft Statistica v12(57).

### 2.4 Magnetic resonance imaging grey matter analysis

A subsample of 61 women also underwent 3 Tesla MRI acquisition postpartum. T1-weighted sequences were acquired on a 3T Ingenia CX scanner, using a 32-channel sensitivity encoding SENSE head coil (T1-weighted MPRAGE sequence: TR 8.00 ms, TE 3.7 ms, FOV = 256 mm, matrix = 256 × 256, in-plane resolution 1 × 1 mm, yielding 182 transversal slices with a thickness of 1 mm) at IRCCS San Raffaele Hospital and on a Philips Achieva D-Stream 3T scanner *(Philips, Best, The Netherlands)* equipped with a 32-channel head coil (T1-weighted turbo field echo SENSE sequence: TR = 8.00 ms, TE = 3.7 ms, FOV = 250 mm (FH) × 240 mm (AP) × 180 mm (RL), voxel size: 1 × 1 × 1 mm^3^, flip angle = 8°) at Fondazione IRCCS Ca’ Granda Ospedale Maggiore Policlinico. T1-weighted sequences were preprocessed through the preprocessing pipeline implemented in CAT12.8 (https://neuro-jena.github.io/cat/) for SPM12 (https://www.fil.ion.ucl.ac.uk/spm/software/spm12/) in Matlab R2016b. The pipeline included segmentation into GM, white matter, and cerebrospinal fluid, bias regularization, non-linear modulation, and normalization to Montreal Neurological institute and Hospital (MNI) space using DARTEL to a 1.5 mm isotropic MNI template. GM images were smoothed with an 8 mm full width at half maximum Gaussian filter. Total intracranial volume (TIV) measurements were obtained. Voxelwise differences in GM volumes across groups were investigated through Voxel Based Morphometry (VBM) analysis, as implemented in SPM12. Analyses were conducted in the general linear model (GLM) framework, entering tissue probability maps, generated during MRI segmentation, as dependent variables. The individual PLS component scores and the single cytokines with VIP>1 were entered as regressors in separate moderation models with PPD status as the moderator. In all analyses, the same covariates were used as in the PLS analyses plus days from delivery to MRI scan and TIV. Threshold masking was set at an absolute value of 0.1. Significance level was set at p_FWE_<0.05 at cluster level with p<0.001 uncorrected at voxel level.

## 3. RESULTS

Clinical and demographic characteristics of the whole sample are resumed in Table 1. Demographics of the neuroimaging subsample can be found in Supplementary Table 1. In agreement with the epidemiology of PPD in the Italian population (40, 41), 30/140 women (21%) scored above threshold on the EPDS. Group comparisons of third trimester and postpartum markers and metabolites can be found in Supplementary Table 2.

**Table 1:** Demographics of the whole sample. **Legend**: HSR (San Raffaele Hospital); PC (IRCCS Ca’ Granda Ospedale Maggiore Policlinico); EPDS (Edinburgh Postnatal Depression Scale)

| Variable | HC (n=110) | PPD (n=30) | t/$\chi^2$ | p-value |
| --- | --- | --- | --- | --- |
| Age | 34.65 ± 4.792 | 35.17 ± 3.196 | 0.691 | 0.492 |
| Education (years) | 16.54 ± 3.294 | 16.54 ± 2.800 | -0.597 | 0.276 |
| Number of previous pregnancies | 0.32 ± 0.578 | 0.52 ± 0.829 | 1.199 | 0.238 |
| First pregnancy (Y/N) | 79/31 | 18/12 | 1.547 | 0.214 |
| Center (HSR/PC) | 63/47 | 21/9 | 1.591 | 0.207 |
| Gestational week at 3<sup>rd</sup> trimester blood sample | 33.96 ± 2.983 | 33.83 ± 3.152 | -0.210 | 0.834 |
| 3<sup>rd</sup> trimester EPDS score | 4.04 ± 3.507 | 8.67 ± 6.357 | 3.431 | 0.002 |
| Days from delivery to blood sampling | 66.37 ± 37.967 | 69.42 ± 42.438 | 0.343 | 0.732 |
| Postpartum EPDS score | 4.07 ± 3.126 | 15.30 ± 4.178 | 16.152 | <0.001 |

### 3.1 Predicting PPD with third trimester circulating levels of signaling molecules

A PLS linear regression with all available third trimester cytokines as regressors defined a significant model predicting PPD, with one significant component (coefficient=0.086) explaining 6.7% of variance: R²X=0.449; R²Y=0.067; Q²=0.032. All nuisance covariates were dismissed as not relevant. 21 circulating molecules had a VIP>1 in predicting PPD, all with positive predictive weights, and all but one confirmed to significantly explain PPD at GLZM regression models (Table 2). Repeating the PLS analysis after excluding the non-significant variables slightly increased the predictive power (R²X=0.565; R²Y=0.074; Q²=0.049), with one significant component predicting 7.4% of variance, without changes in the strength and direction of effect of the single variables.

**Table 2:** Signaling cytokine and chemokine molecules significantly predicting postpartum depression at partial least squared regression, and at single generalized linear models. **Legend:** PLS (partial least squares regression); GLZM (generalized linear model); VIP (variable importance in projection); w (weight); LR χ^2^ (Likelihood Ratio Chi-Square test); p (p-value)

|  |  | PLS regression |  | GLZM |  |
| --- | --- | --- | --- | --- | --- |
| Rank | Variable | VIP | w | LR$\chi^2$ | p |
| Circulating levels at the third trimester |  |  |  |  |  |
| 1 | MIP1α (CCL3) | 1.738 | 0.253 | 9.906 | 0.0016 |
| 2 | TNF-β (LT- $\alpha$) | 1.721 | 0.251 | 9.620 | 0.0019 |
| 3 | IL-10 | 1.654 | 0.241 | 7.474 | 0.0063 |
| 4 | G-CSF | 1.619 | 0.236 | 8.137 | 0.0043 |
| 5 | M-CSF | 1.572 | 0.229 | 7.717 | 0.0055 |
| 6 | IL-18 | 1.502 | 0.219 | 7.469 | 0.0063 |
| 7 | IL-7 | 1.486 | 0.217 | 6.993 | 0.0082 |
| 8 | GRO $\alpha$ (CXCL1) | 1.477 | 0.215 | 6.945 | 0.0084 |
| 9 | HGF | 1.420 | 0.207 | 6.713 | 0.0096 |
| 10 | IL-16 | 1.378 | 0.201 | 6.309 | 0.0120 |
| 11 | TNF-α | 1.360 | 0.198 | 6.068 | 0.0137 |
| 12 | SCF | 1.358 | 0.198 | 6.215 | 0.0127 |
| 13 | β-NGF | 1.281 | 0.187 | 4.779 | 0.0288 |
| 14 | MIP-1β | 1.210 | 0.176 | 5.147 | 0.0233 |
| 15 | MIF | 1.184 | 0.173 | 4.606 | 0.0319 |
| 16 | SDF1 $\alpha$ | 1.175 | 0.171 | 4.752 | 0.0293 |
| 17 | TRAIL | 1.144 | 0.167 | 4.326 | 0.0375 |
| 18 | IL-2 | 1.126 | 0.164 | 4.049 | 0.0442 |
| 19 | IL1ra | 1.105 | 0.161 | 4.044 | 0.0443 |
| 20 | LIF | 1.082 | 0.158 | 3.939 | 0.0472 |
| 21 | IL1α | 1.019 | 0.149 | 3.443 | 0.0635 |
| Circulating levels postpartum |  |  |  |  |  |
| 1 | IL-7 | 1.842 | 0.461 | 7.890 | 0.0050 |
| 2 | TRAIL | 1.719 | 0.430 | 6.693 | 0.0010 |
| 3 | MIP1α (CCL3) | 1.677 | 0.419 | 6.745 | 0.0010 |
| 4 | IL-18 | 1.317 | 0.329 | 4.122 | 0.0423 |
| 5 | TNF-β (LT- $\alpha$) | 1.190 | 0.298 | 3.482 | 0.0620 |
| 6 | IL1ra | 1.002 | 0.251 | 2.491 | 0.1146 |

Testing the individual cytokines PLS component in a CART algorithm yielded a classification matrix with high predictive values and overall accuracy of 87.94% in identifying PPD at an estimated a priori disease prevalence 20% (Table 3).

**Table 3:** Outcome of classification and regression tree (CART) algorithm for third trimester cytokines partial least squares regression (PLS) component: confusion matrix and predictive values. **Legend:** (*) The result is influenced by the prevalence of the condition in the population.

|  | Actually Positive | Actually Negative |
| --- | --- | --- |
| **Predicted Positive** | 13 | 1 |
| **Predicted Negative** | 17 | 109 |
| **Statistic** | **Value** | **95% CI** |
| Sensitivity | 43.33% | 25.46% to 62.57% |
| Specificity | 99.09% | 95.04% to 99.98% |
| Positive Predictive Value (*) | 92.26% | 61.88% to 98.87% |
| Negative Predictive Value (*) | 87.49% | 83.64% to 90.54% |
| Accuracy (*) | 87.94% | 81.37% to 92.83% |

KP metabolites also predicted PPD. Significant effects were detected for the KYNA/QUIN ratio (VIP=1.887, w=-0.629), with KYNA protecting against PPD (VIP=1.646, w=-0.549), and QUIN fostering PPD (VIP=1.38, w=0.379). The other metabolites and ratios were discarded as not significant. The PLS model could significantly explain 3.2% of variance: coefficient=0.184; R²X=0.636; R²Y=0.032; Q²=0.009. However, testing the effect of these factors in a GLZM regression only gave marginal effects for the KYNA/QUIN ratio (LRχ^2^=3.333, p=0.0679), suggesting that KP metabolites can predict PPD through their multivariate signature rather than via isolated effects on single metabolites.

Adding the PLS component for KP metabolites to that for cytokines in a CART algorithm to predict PPD at a prevalence of 20%, substantially increased sensitivity, also maintaining a high overall accuracy of prediction at 93.27% (Table 4).

**Table 4:**
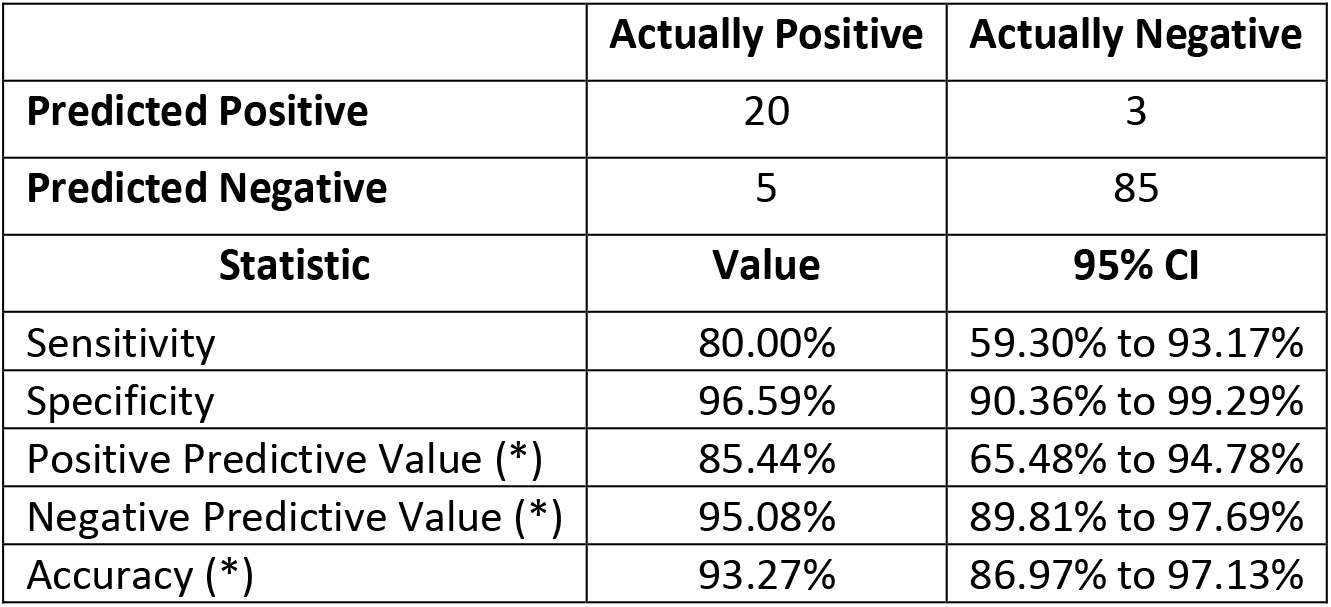
Outcome of classification and regression tree (CART) algorithm for third trimester cytokines partial least squares regression (PLS) component and kynurenine metabolites PLS component: confusion matrix and predictive values. **Legend:** (*) The result is influenced by the prevalence of the condition in the population.

|  | Actually Positive | Actually Negative |
| --- | --- | --- |
| **Predicted Positive** | 20 | 3 |
| **Predicted Negative** | 5 | 85 |
| **Statistic** | **Value** | **95% CI** |
| Sensitivity | 80.00% | 59.30% to 93.17% |
| Specificity | 96.59% | 90.36% to 99.29% |
| Positive Predictive Value (*) | 85.44% | 65.48% to 94.78% |
| Negative Predictive Value (*) | 95.08% | 89.81% to 97.69% |
| Accuracy (*) | 93.27% | 86.97% to 97.13% |

### 3.2 Association of postpartum circulating levels of signaling molecules with PPD

A PLS model including postpartum levels of the cytokines and chemokines that significantly predicted PPD when measured at the third trimester failed to predict PPD, despite explaining 5.2% of variance (R²X=0.643; R²Y=0.052; Q²=-0.039). Six analytes, however, showed a VIP>1, four of which also showed a significant effect when entered as factors in a GLZM regression (Table 2).

A PLS model with postpartum KP metabolites which significantly predicted PPD, when measured at the third trimester, could still significantly explain 1.2% of variance and predict PPD: coefficient=0.113; R²X=0.648; R²Y=0.012; Q²=0.004. Selected variables were the KYNA/QUIN ratio (VIP=1.318, w=-0.761), and KYNA (VIP=1.109, w=-0.640), protecting against PPD. However, effects were not significant when tested at GLZM regression.

### 3.3 Association of third trimester circulating levels of signaling molecules with antepartum depression

116 participants completed the EPDS at the third trimester, among which 15 (12.9%) scored above the threshold. Of these, 6/15 (40%) then scored below threshold postpartum, while 15/101 (14.9%) of the women below threshold at the third trimester then developed PPD. Despite these substantial changes in depression status, antepartum depression remained significantly associated with PPD (χ^2^=16.22, p<0.0001).

A PLS model with third trimester analytes as regressors defined a significantly predicted antepartum depression, with one significant component (coefficient=0.076) explaining 4.9% of variance (R²X=0.494; R²Y=0.053; Q²=0.029). 19 circulating molecules had a VIP>1, all with positive predictive weights. Twelve of these variables also predicted PPD, suggesting that the association between specific cytokines and depression preceded delivery. However, only the 6 top ranking molecules confirmed to significantly explain antepartum depression at GLZM regression models. Results are detailed in Supplementary Table 3.

### 3.4 Brain imaging

#### 3.4.1 Third trimester cytokine and chemokine interactions

There was a significant interaction between the component scores from the PLS using third trimester cytokines and chemokines with a VIP>1 and PPD status on a cluster including left occipital regions. Post hoc analysis revealed a positive association between the component and the extracted cluster in HC and a negative association in PPD women. Significant interactions were also found for the single cytokines IL-10, IL-7, SCF, β-NGF, MIP-1β, IL-2, and IL-1ra. Most of the cytokines followed a similar pattern as the component for directionality in largely overlapping brain regions, besides IL-10 which had an opposite association of being negatively associated to GMV in HC and positively associated in PPD women. Results of the VBM and SPSS analyses can be found in Table 5.

**Table 5:** Voxel based morphometry and SPSS post-hoc analyses of third trimester cytokine and chemokine factors. **Legend:** VBM (Voxel based morphometry); SPSS (statistical package for the social sciences); HC (healthy controls); PPD (postpartum depression)

| Marker | VBM contrast | Cluster size | Cluster level significance | Peak level significance | SPSS interaction | HC effect | PPD effect | Brain areas of cluster |
| --- | --- | --- | --- | --- | --- | --- | --- | --- |
| PLS component score | (HC>PPD) | 877 | pFWE=0.031 | pFWE=0.164 | $\beta$=-0.0091, p<.001 | $\beta$=0.0032, p=.003 | $\beta$=-0.0059, p<.001 | L occipital pole, L calcarine cortex, L inferior, superior, and middle occipital gyri |
| IL-10 | (HC<PPD) | 345 | pFWE=0.315 | pFWE=0.029 | $\beta$=0.0042, p<.001 | $\beta$=-0.002, p<.001 | $\beta$=0.0018, p=.004 | L fusiform gyrus, L inferior temporal gyrus, L parahippocampal gyrus |
| IL-7<br>Cluster 1 | (HC>PPD) | 976 | pFWE=0.020 | pFWE=0.246 | $\beta=-0.0004$,<br>$p<.001$ | $\beta=0.0002$,<br>$p<.001$ | $\beta=-0.0002$,<br>$p<.001$ | L postcentral and<br>precentral gyri, L<br>superior parietal<br>lobule |
| IL-7<br>Cluster 2 | (HC>PPD) | 616 | pFWE=0.091 | pFWE=0.010 | $\beta=-0.0004$,<br>$p<.001$ | $\beta=0.0002$,<br>$p<.001$ | $\beta=-0.0002$,<br>$p=.001$ | L occipital pole, L<br>calcarine cortex, L<br>inferior and<br>superior and<br>middle occipital<br>gyri |
| SCF | (HC>PPD) | 1551 | pFWE=0.003 | pFWE=0.044 | $\beta=-0.0012$,<br>$p<.001$ | $\beta=0.0004$,<br>$p=.002$ | $\beta=-0.0008$,<br>$p<.001$ | L inferior occipital<br>gyrus |
| β-NGF | (HC>PPD) | 1188 | pFWE=0.008 | pFWE=0.166 | $\beta=-0.0128$,<br>$p<.001$ | $\beta=.0023$,<br>$p=.1761$ | $\beta=-0.0105$,<br>$p<.001$ | R middle and<br>superior temporal<br>gyri, R angular<br>gyrus |
| MIP-1β | (HC>PPD) | 1269 | pFWE=0.007 | pFWE=0.097 | $\beta=-0.0006$,<br>$p<.001$ | $\beta=0.0002$,<br>$p=.004$ | $\beta=-0.0004$,<br>$p<.001$ | L calc calcarine<br>cortex, L occipital<br>pole, L cuneus, and<br>L lingual gyrus |
| IL-2<br>Cluster 1 | (HC>PPD) | 381 | pFWE=0.269 | pFWE=0.021 | $\beta=-0.0043$,<br>$p<.001$ | $\beta=0.0016$,<br>$p=.022$ | $\beta=-0.0027$,<br>$p<.001$ | L superior frontal<br>gyrus medial<br>segment, L<br>supplementary<br>motor cortex L<br>middle and anterior<br>cingulate gyrus,<br>and L superior<br>frontal gyrus |
| IL-2<br>Cluster 2 | (HC>PPD) | 347 | pFWE=0.316 | pFWE=0.041 | $\beta=-0.0039$,<br>$p<.001$ | $\beta=0.0019$,<br>$p<.001$ | $\beta=-0.0020$,<br>$p=.001$ | L occipital pole, L<br>calcarine cortex, L<br>superior, inferior,<br>and middle<br>occipital gyri |
| IL1ra | (HC>PPD) | 696 | pFWE=0.065 | pFWE=0.023 | $\beta=-0.0001$,<br>$p<.001$ | $\beta<.001$,<br>$p=.012$ | $\beta<-.001$,<br>$p<.001$ | L inferior occipital<br>gyrus |

#### 3.4.2 Third trimester kynurenine pathway interactions

There was a significant interaction between the component scores from the PLS using third trimester KP metabolites with a VIP>1 and PPD status on a cluster of the right precentral and postcentral gyri. Post hoc analysis revealed no association between the component and the extracted cluster in HC and a positive association in PPD women. A significant interaction was also found for KYNA/QUIN ratio following a similar pattern as the component in the same brain cluster. Results of the VBM and SPSS analyses can be found in Table 6.

**Table 6:**
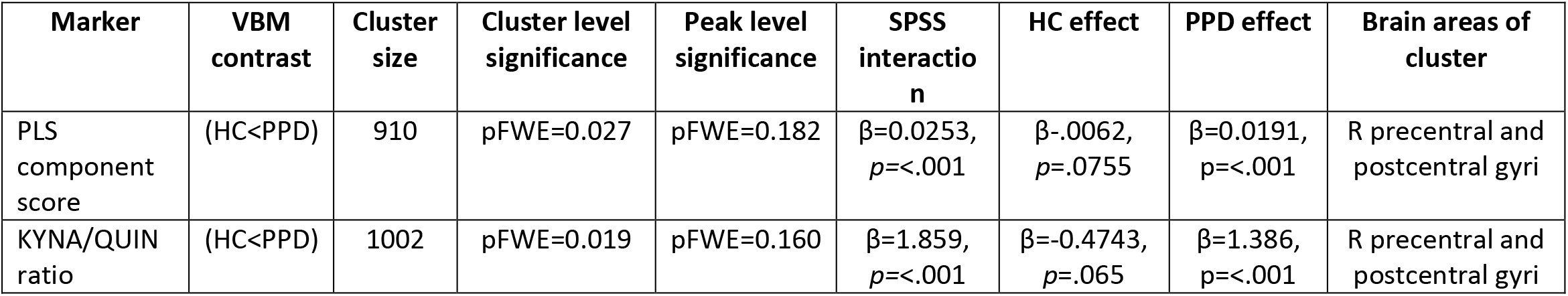
Voxel based morphometry and SPSS post-hoc analyses of third trimester kynurenine pathway factors. **Legend:** VBM (Voxel based morphometry); SPSS (statistical package for the social sciences); HC (healthy controls); PPD (postpartum depression)

## 4. DISCUSSION

We provide evidence that immune-inflammatory and KP alterations during late pregnancy can predict PPD and have different associations with postpartum brain morphology based on PPD status.

The biological mechanisms underlying these associations are likely multifactorial, yet the placenta may plausibly play a central role in regulating the immune adaptations that characterize pregnancy. The placenta functions as an immune-modulating organ that regulates the immune responses of cells present both at the implantation site and systemically, to promote immune tolerance and to ensure a maximal protection for the mother and fetus (58). Disruption of these coordinated adaptations may contribute to both obstetric complications and vulnerability to depression (59, 60).

The cytokines that predicted PPD play specific roles in pregnancy, either in fostering immune tolerance at the maternal-fetal interface, or in promoting a pro-inflammatory milieu (Table 7). Given that many of these molecules are produced at the placental and fetal level and decline sharply after delivery, this pattern is consistent with a placental contribution. A similar placental contribution can be hypothesized for KYN metabolites, given that severe antenatal depression has been associated with lower placental levels of enzymes that degrade QUIN, thus leading to higher QUIN/KYNA ratio (61), a feature we showed to predict PPD. These effects could be potentially relevant to predict and prevent PPD in clinical settings. When introducing the PLS component scores in a CART algorithm, we observed a high predictive accuracy, with a high specificity, meaning that when PPD is predicted, it has a high probability to be observed. In medical settings, this could foster special care and possible preventative intervention once the specificity for the novel panel of predictors has been confirmed in independent studies.

**Table 7:**
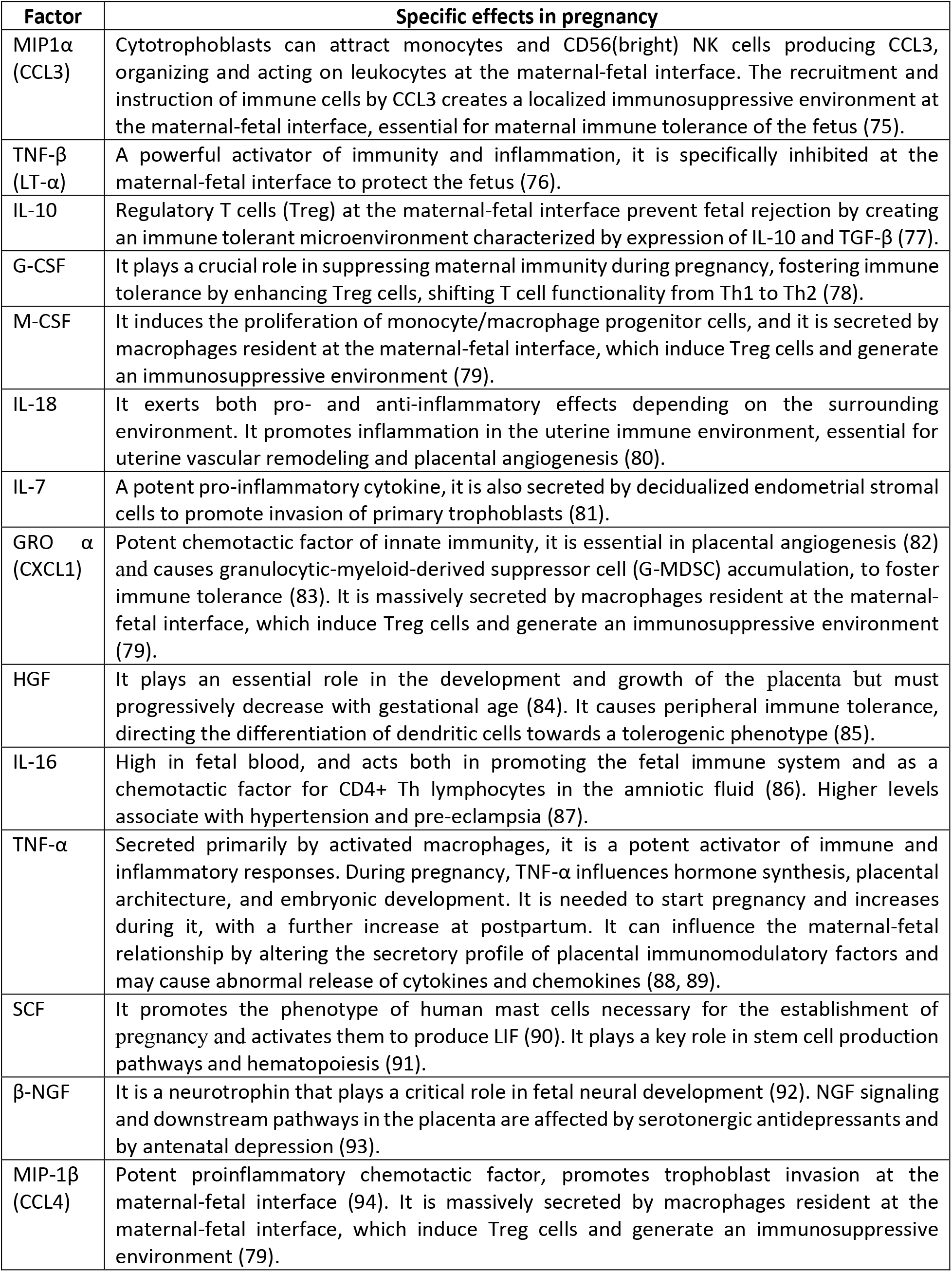
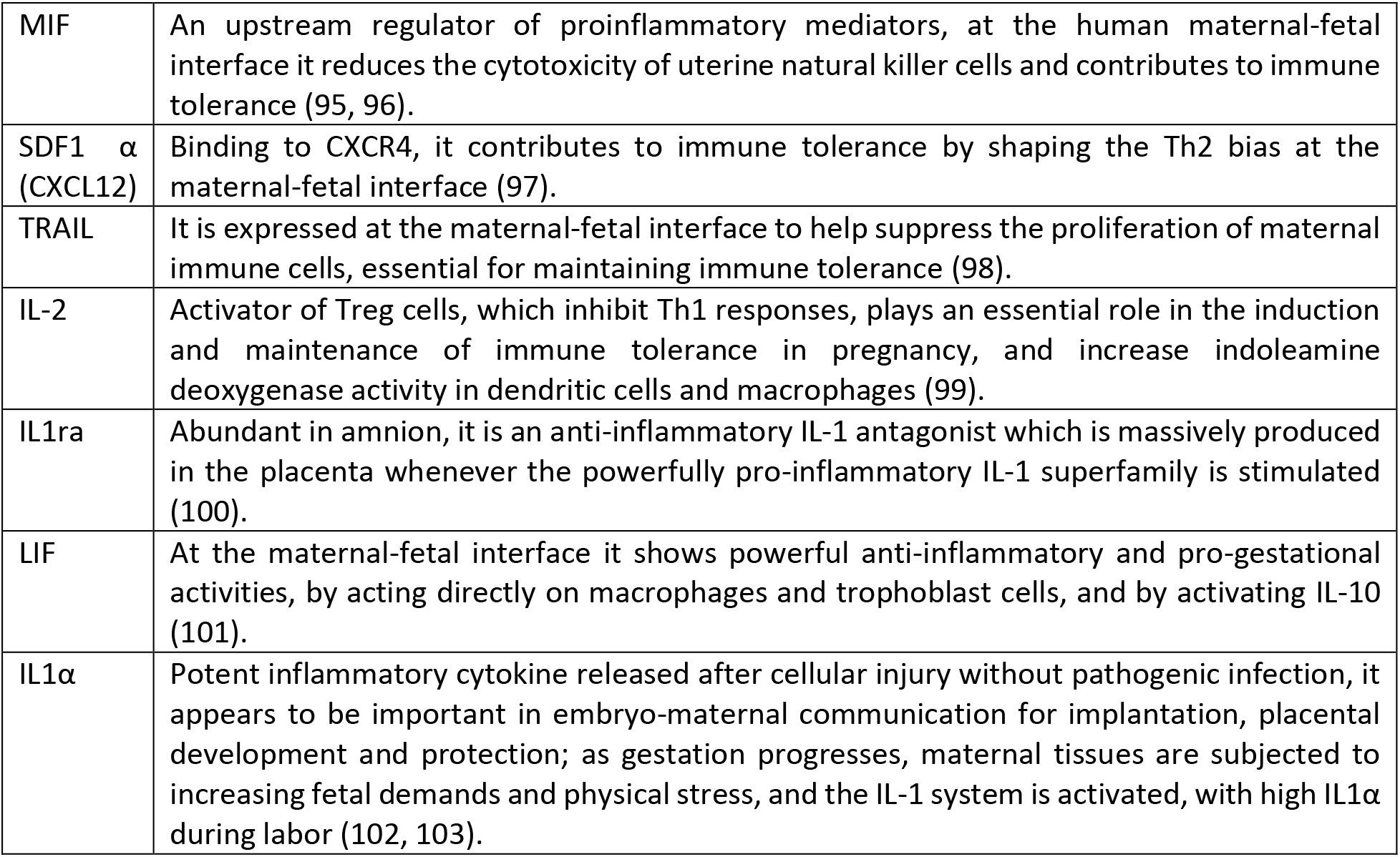
Specific pregnancy related effects of immune-inflammatory signaling molecules, which significantly predicted the occurrence of postpartum depression when measured at the 3^rd^ trimester.

It can be surmised that once produced and released into the bloodstream, these molecules can act on the multiple targets linking brain function and immune-inflammatory setpoints in depression, including specific brain effects on monoaminergic and glutamatergic neurotransmission, neurovascular coupling, and general brain homeostasis and microstructure (53, 62–64). In our neuroimaging analyses we discovered that the relationship between third trimester inflammation and postpartum brain morphology differed according to PPD status. The inflammatory component derived from third trimester cytokines and chemokines showed a significant interaction with PPD in a large cluster encompassing occipital and calcarine regions. In particular we found that third trimester inflammation had a negative association with the cluster in PPD women, yet had a positive association in women who did not develop PPD. These patterns suggest that the neuroanatomical consequences of inflammatory signaling during late pregnancy may depend on an individual’s vulnerability to PPD.

Despite their diverse biological functions, several of the individual cytokines, including IL-7, SCF, MIP1β, IL-2, and IL-1ra, demonstrated remarkably similar interaction patterns, which could suggest that the observed effects may reflect broader immune-related processes rather than the influence of isolated inflammatory molecules. Pregnancy is also accompanied by profound neural adaptations, many of which are thought to support maternal behaviors and facilitate adaptation to infant care (28, 32). In this context, inflammatory signaling may in part aid in physiological neuroplastic processes in some women, while contributing to maladaptive structural changes in others that are more sensitive to inflammation. Indeed, previous studies have found that inflammatory markers have a negative effect on the brain in mood disorders also known to be sensitive to neuroinflammation (65–67).

Many of the significant cytokine-related clusters were located within occipital, calcarine, cuneal, and lingual regions. Although these areas are classically associated with visual processing, they are also thought to be important for emotional processing of sensory information (68). Studies in PPD have found alternations in visual network connections among the cognitive control network and default mode network which could interrupt emotional processing of infant sensory information and lead to increased self-referential bias typical of depressive disorders (69). In fact, emotional processing differences have been found in PPD compared with healthy mothers with one study finding increased sustained attention on negative facial expressions from infants and adults and reduced processing on adult happy faces compared to healthy mothers and non-mothers who preferred happy faces (70).

A partially distinct pattern emerged for third trimester KP measures. Both the KP component and the KYNA/QUIN ratio showed significant interactions with PPD within overlapping clusters encompassing the right precentral and postcentral gyri. In contrast to the cytokine findings, KP measures were positively associated with GMV only among women who developed PPD, whereas no significant association was observed in HC. The KP links immune activation with neuroplasticity and glutamatergic neurotransmission (71, 72). Consistent with the neuroprotective properties of KYNA and the excitotoxic effects of QUIN, the KYNA/QUIN ratio is often used as an index of the balance between protective and potentially neurotoxic kynurenine metabolism (73). The precentral and postcentral gyri form the primary sensorimotor cortex and play a central role in integrating sensory and motor information (74). Therefore, given the substantial sensory and behavioral adaptations that accompany pregnancy and early motherhood, future studies should investigate whether alterations in kynurenine metabolism are linked to changes in sensorimotor processing relevant to maternal adaptation.

In conclusion, the present findings suggest that immune-inflammatory and KP alterations during late pregnancy are associated with both vulnerability to PPD and differences in postpartum brain morphology. Our study is the first to capture these complex predictive patterns through a solid multivariate framework. The predictive value of third-trimester biomarkers, together with their associations with grey matter volume measured after delivery, supports the hypothesis that biological processes occurring during pregnancy contribute to neurobiological pathways relevant to PPD and could potentially help to identify women at increased risk. However, several limitations should be considered. First, PPD status was determined based on a self-rated questionnaire rather than a clinical diagnosis. Second, given the relatively small neuroimaging sample and the large number of biomarkers examined, individual marker findings should be interpreted cautiously until replicated in independent cohorts. Third, MRI was only acquired postpartum therefore limiting the causal inference of our findings. Future longitudinal studies integrating placental, immune, metabolic, and neuroimaging measures will be important to clarify the mechanisms linking pregnancy-related biological adaptations to maternal mental health and to evaluate the potential clinical utility of these biomarkers for the early identification of women at increased risk for postpartum depression.

**Figure 1:**
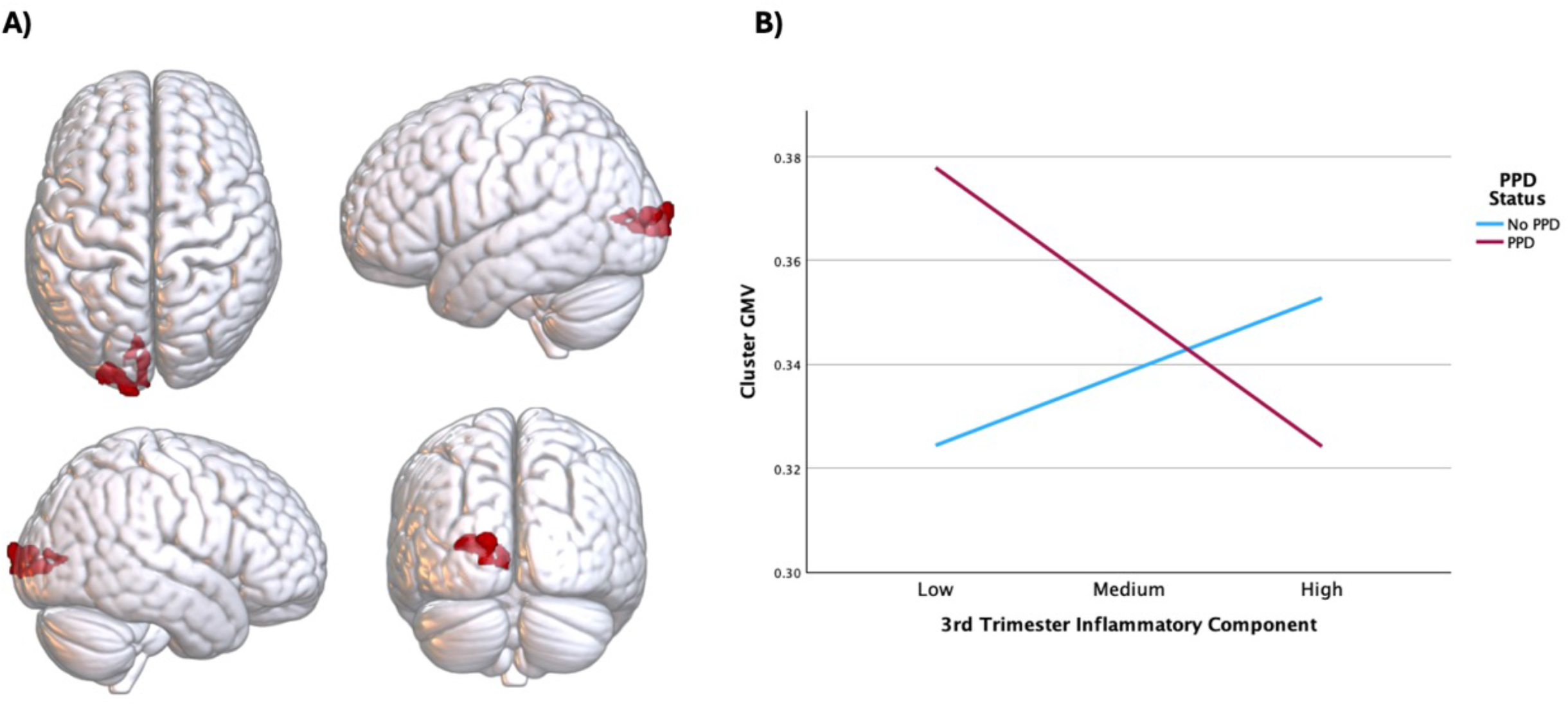
**A)** Occipital cortex cluster identified in the voxel-based morphometry analysis consisting of the L occipital pole, L calcarine cortex, L inferior, superior, and middle occipital gyri to be differentially affected by the third trimester cytokine and chemokine inflammatory PLS component based on postpartum depression (PPD) status. **B)** Moderation analysis showing a significant interaction between the third trimester cytokine and chemokine inflammatory component and PPD status in the extracted occipital cortex cluster with a positive effect in no PPD women and a negative effect in PPD women

**Figure 2:**
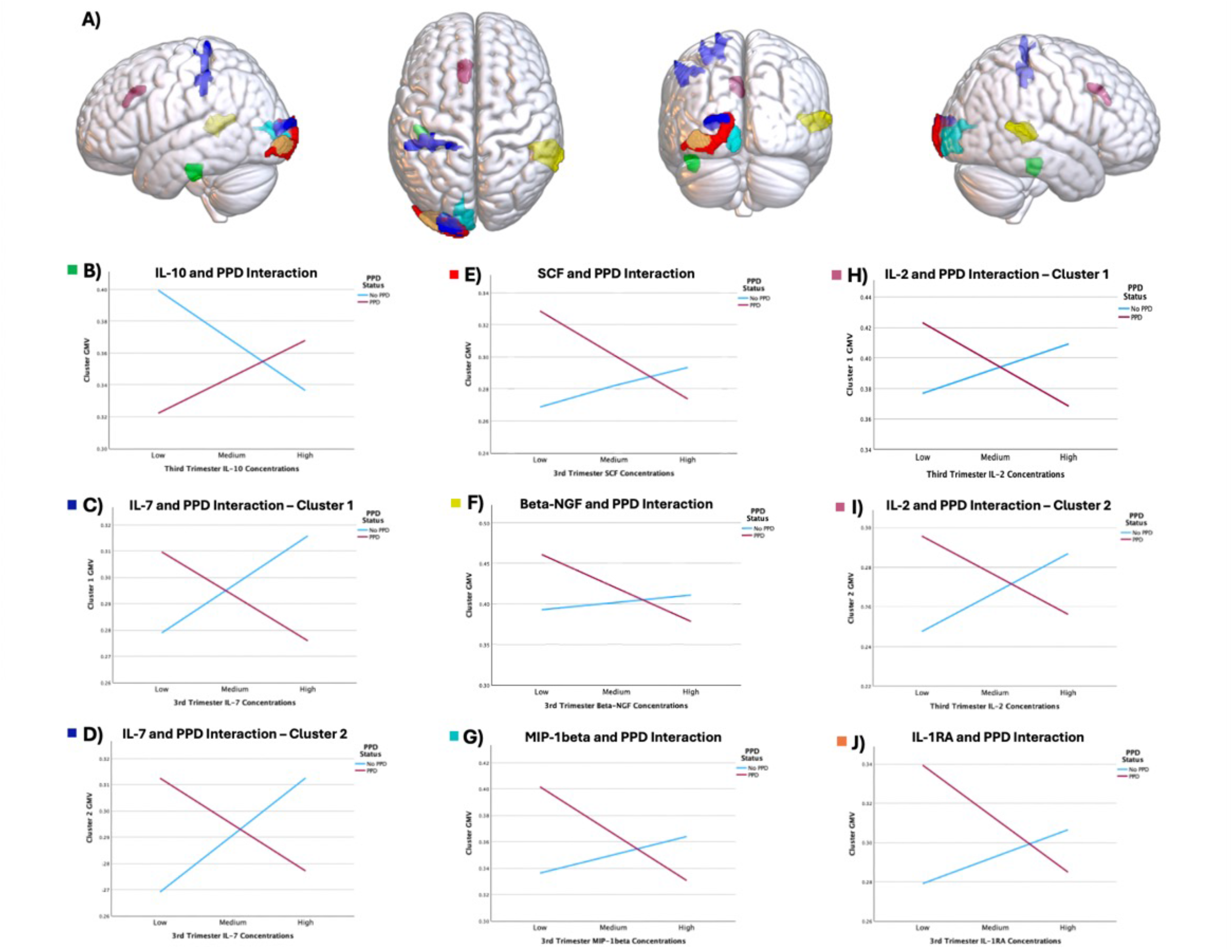
**A)** Clusters identified in the voxel based morphometry analysis for the individual cytokine moderation analysis between PPD and IL-10 (green), IL7 (blue), SCF (red), beta-NGF (yellow), MIP-1beta (teal), IL-7 (magenta), and IL-1ra (orange). **B)** Moderation analysis showing a significant interaction between third trimester IL-10 and PPD in an extracted cluster encompassing the L fusiform gyrus, L inferior temporal gyrus, L cerebral W matter, and L parahippocampal gyrus. **C)** Moderation analysis showing a significant interaction between third trimester IL-7 and PPD in the first extracted cluster encompassing the L postcentral gyrus, L cerebral W matter, L precentral gyrus, and L superior parietal lobule. **D)** Moderation analysis showing a significant interaction between third trimester IL-7 and PPD in the second extracted cluster encompassing the L cerebral W matter, L occipital pole, L calcarine cortex, and L inferior and superior and middle occipital gyri. **E)** Moderation analysis showing a significant interaction between third trimester SCF and PPD in the extracted cluster encompassing the L inferior occipital gyrus. **F)** Moderation analysis showing a significant interaction between third trimester beta-NGF and PPD in the extracted cluster encompassing the R middle and superior temporal gyri, R cerebral W matter, R angular gyrus. **G)** Moderation analysis showing a significant interaction between third trimester MIP-1beta and PPD in the extracted cluster encompassing the L calc calcarine cortex, L occipital pole, L cerebral W matter, L cuneus, and L lingual gyrus. **H)** Moderation analysis showing a significant interaction between third trimester IL-2 and PPD in the first extracted cluster encompassing the L cerebral W matter, L superior frontal gyrus medial segment, L supplementary motor cortex L middle and anterior cingulate gyrus, and L superior frontal gyrus. **I)** Moderation analysis showing a significant interaction between third trimester IL-2 and PPD in the second extracted cluster encompassing the L cerebral W matter, L occipital pole, L calcarine cortex, L superior and inferior and middle occipital gyrus. **J)** Moderation analysis showing a significant interaction between third trimester MIP-1beta and PPD in the extracted cluster encompassing the L inferior occipital gyrus.

**Figure 3:**
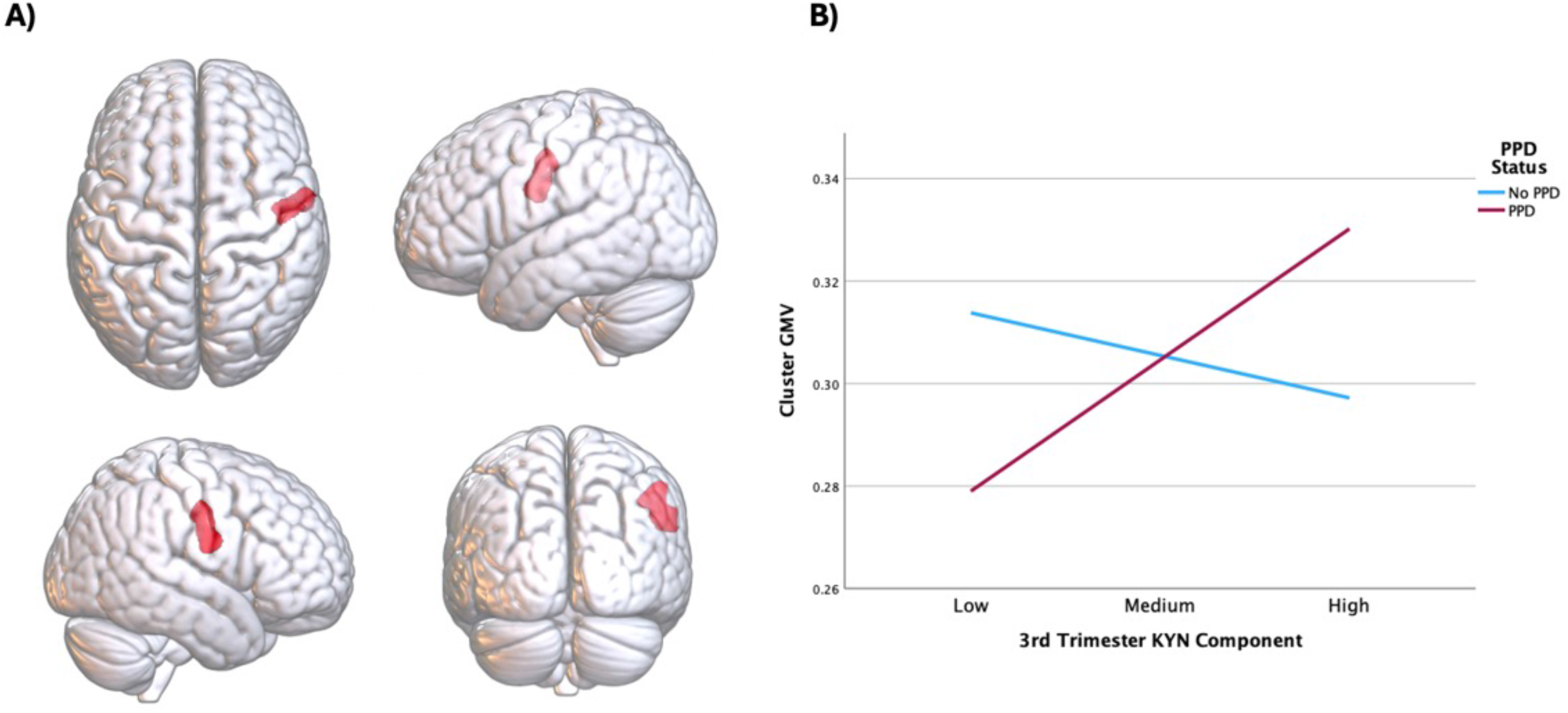
**A)** Right cluster identified in the voxel-based morphometry analysis consisting of the right precentral and postcentral gyri to be differentially affected by the third trimester kynurenine pathway component based on PPD status. **B)** Moderation analysis showing a significant interaction between the third trimester kynurenine metabolites component and PPD status in the extracted pre/postcentral gyri cluster with no effect in HC women and positive effect in PPD women

**Figure 4:**
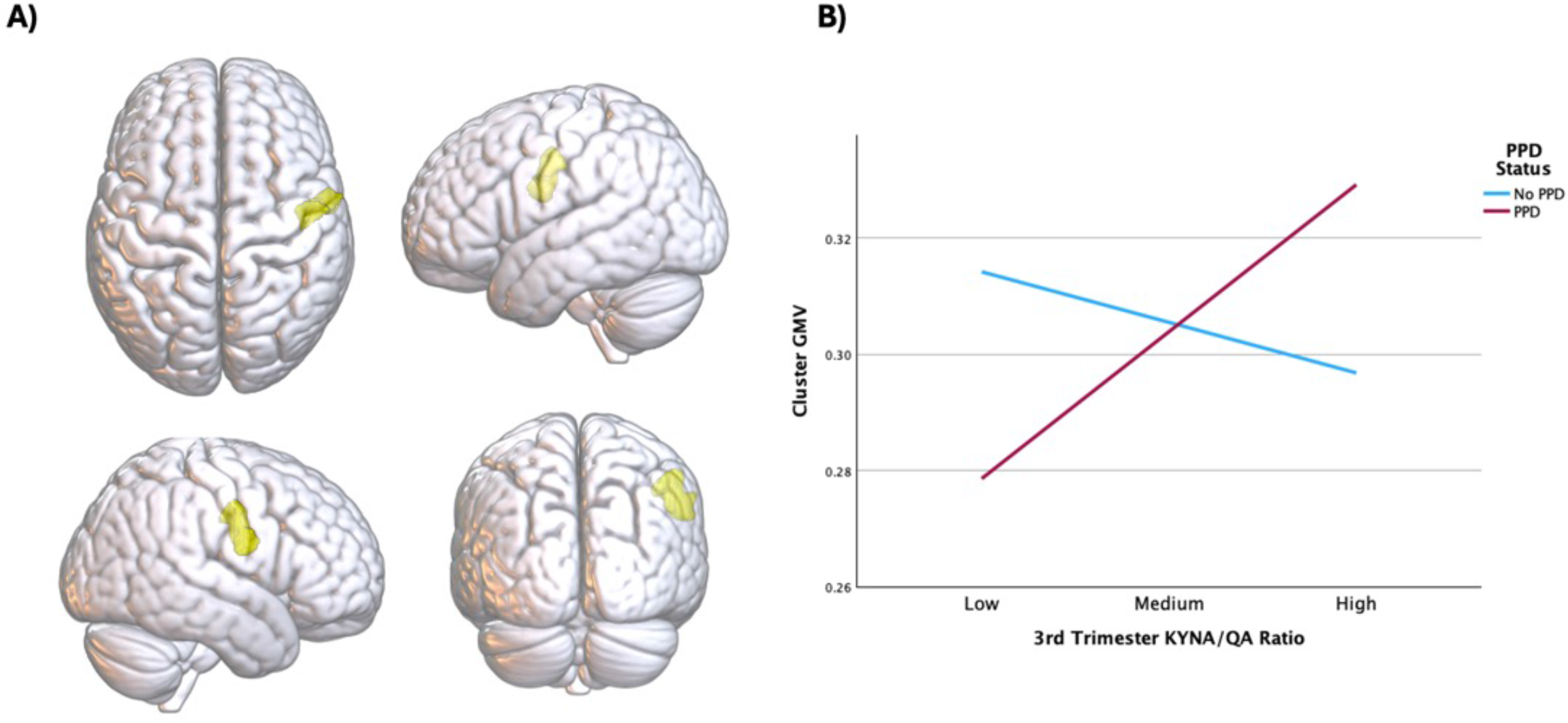
**A)** Right cluster identified in the voxel-based morphometry analysis consisting of the right precentral and postcentral gyri to be differentially affected by the third trimester kynurenic acid (KYNA) and quinolinic acid (QUIN) ratio based on PPD status. **B)** Moderation analysis showing a significant interaction between the third trimester kynurenic acid (KYNA) quinolinic acid (QUIN) ratio and PPD status in the extracted pre/postcentral gyri cluster with no effect in no PPD women and positive effect in PPD women

## Data Availability

All data produced in the present study are available upon reasonable request to the authors.

## Funding source

This work was supported by the Italian Ministry of Health, project RF-2019-12371066 (FB, MCM)

## Author contributions

FB, YAH, EMTM, and PB designed the study; HKS, CM, MCM, AM, EM, LS, ET, PE, NP, GC, EMTM, EA, SS, CL, SN, SDA, SC, SP, MO, and PB organized and performed recruitment and data processing; SS, CL, SN, SDA performed lab analysis; YAH, HKS, CM, and FB performed statistical analysis; YAH and FB wrote the first draft, and all authors edited and approved the final version of the submitted manuscript.

## Contributors/Collaborators

The Milano PPDINFLAME Study Group includes:

Beatrice Arosio, Simona Boito, Irene Cetin, Giuseppe Delvecchio, Valentina Di Mattei, Virginia Favalli, Adele Ferro, Francesca Gatti, Serena Giomi, Diletta Iraci, Alessandra Marano, Francesca Milano, Gaia Perego, Nicola Persico, Fabio Maria Triulzi, and Zaffaella Zanardi.

## Prior presentations

A preliminary version of this study was submitted to the 39^th^ European College of Neuropsychopharmacology (ECNP) as an abstract for a poster presentation.

## DISCLOSURES

The authors report no biomedical financial interests or potential conflicts of interest.

---

**Supplementary Table 1:** Demographics of the voxel based morphometry analysis sample.

| Variable | HC (n=46) | PPD (n=15) | t/$\chi^2$ | p-value |
| --- | --- | --- | --- | --- |
| Age | 34.39 ± 5.157 | 35.60 ± 3.562 | 0.842 | 0.403 |
| Education (years) | 16.58 ± 3.714 | 16.20 ± 2.731 | -0.362 | 0.719 |
| Number of Previous pregnancies | 0.33 ± 0.560 | 0.47 ± 0.743 | 0.777 | 0.440 |
| First pregnancy (Y/N) | 33/13 | 10/5 | 1.40 | 0.708 |
| Center (HSR/PC) | 26/20 | 9/6 | 0.56 | 0.813 |
| Gestational week at 3<sup>rd</sup> trimester blood sample | 33.61 ± 2.206 | 34.00 ± 3.295 | 0.430 | 0.672 |
| 3<sup>rd</sup> Trimester EPDS score | 3.73 ± 3.421 | 6.23 ± 5.183 | 1.620 | 0.125 |
| Days from delivery to blood sampling | 57.31 ± 28.994 | 73.07 ± 40.777 | 1.639 | 0.107 |
| Postpartum EPDS score | 4.07 ± 2.999 | 16.00 ± 4.826 | 11.405 | <0.001 |
| Days from delivery to MRI | 80.20 ± 50.263 | 103.93 ± 81.351 | 1.350 | 0.182 |
| TIV | 1359.13 ± 104.86 | 1355.47 ± 128.20 | -0.111 | 0.912 |

**Legend:** HSR (San Raffaele Hospital); PC (IRCCS Ca' Granda Ospedale Maggiore Policlinico);
EPDS (Edinburgh Postnatal Depression Scale); MRI (Magnetic resonance imaging); TIV
(Total intracranial volume)

**Supplementary Table 2:** Group comparison of third trimester and postpartum inflammatory markers and kynurenine pathway metabolites between women who developed postpartum depression (PPD) and those who did not (HC)

|  | Third trimester |  | Post partum |  | F, p | Groups<br>(PPD vs HC) |  |
| --- | --- | --- | --- | --- | --- | --- | --- |
|  | PPD | HC | PPD | HC |  | Third trimester | Postpartum |
| IL-2Rα | 60.37<br>(32.51) | 50.2<br>(35.39) | 57.93<br>(29.70) | 47.12<br>(28.88) | 1.86, p=0.176 | p=0.125 | p=0.144 |
| MIG | 219.05<br>(106.32) | 237.70<br>(164.96) | 220.42<br>(100.33) | 335.20<br>(429.05) | 1.00, p=0.319 | p=0.559 | p=0.196 |
| MIP-1β | 341.28<br>(82.62) | 302.24<br>(79.96) | 293.79<br>(90.76) | 282.30<br>(72.39) | 5.495, p=0.021 | p=0.020 | p=0.506 |
| IL-6 | 10.59<br>(5.38) | 8.64<br>(5.36) | 11.99<br>(2.83) | 10.34<br>(4.68) | 0.189, p=0.666 | p=0.176 | p=0.229 |
| IFN-α2 | 30.92<br>(12.88) | 28.81<br>(15.38) | 25.26<br>(14.72) | 25.60<br>(16.40) | 1.74, p=0.189 | p=0.5 | p=0.927 |
| IFN-gamma | 58.03<br>(55.67) | 30.20<br>(39.51) | 51.68<br>(48.76) | 34.08<br>(43.99) | 4.28, p=0.042 | p=0.009 | p=0.126 |
| SDF-1α | 2213.53<br>(899.44) | 1828.55<br>(806.39) | 2302.54<br>(1120.22) | 1918.88<br>(1026.33) | 0.45, p=0.502 | p=0.025 | p=0.107 |
| **IL-1ra** | 824.47<br>(582.32) | 586.48<br>(532.87) | 676.68<br>(513.73) | 496.98<br>(475.55) | 0.287, p=0.593 | p=0.035 | p=0.103 |
| **MCP-3** | 6.18<br>(3.65) | 5.68<br>(4.05) | 3.91<br>(2.56) | 4.99<br>(3.88) | 3.72, p=0.056 | p=0.548 | p=0.216 |
| **IL-16** | 125.35<br>(138.42) | 87.79<br>(37.91) | 122.09<br>(118.71) | 99.21<br>(58.91) | 2.86, p=0.093 | p=0.009 | p=0.172 |
| **IL-12**<br>**(p40)** | 188.69<br>(188.69) | 166.04<br>(226.578) | 106.23<br>(88.09) | 141.28<br>(219.97) | 2.62, p=0.109 | p=0.628 | p=0.476 |
| **LIF** | 121.97<br>(75.24) | 95.58<br>(56.94) | 99.02<br>(66.78) | 88.54<br>(58.58) | 5.38, p=0.022 | p=0.038 | p=0.444 |
| **TNF-β** | 655.40<br>(216.80) | 529.81<br>(168.92) | 593.62<br>(229.33) | 515.84<br>(158.61) | 6.18, p=0.014 | p=<.001 | p=0.050 |
| **IL-5** | 188.26<br>(126.33) | 177.34<br>(104.94) | 269.12<br>(.) | 122.38<br>(105.66) | 0.90, p=0.783 | p=0.847 | P=0.242 |
| **GM-CSF** | 9.83<br>(7.27) | 9.30<br>(6.85) |  | 10.68<br>(7.77) | 0.75, p=0.401 | p=0.843 | p=0.302 |
| **MIF** | 1711.96<br>(1017.28) | 1276.58<br>(892.58) | 1152.34<br>(666.74) | 1207.54<br>(956.57) | 5.29, p=0.023 | p=0.023 | p=0.789 |
| **TNF-α** | 169.24<br>(83.49) | 134.17<br>(58.35) | 134.37<br>(71.12) | 120.91<br>(54.47) | 6.98, p=0.009 | p=0.009 | p=0.333 |
| **RANTS** | 27859.99<br>(37547.21) | 21098.59<br>(46862.21) | 11432.91<br>(10604.30) | 15703.39<br>(25835.81) | .000, p=0.996 | p=0.728 | p=0.428 |
| **IL-2** | 13.78<br>(10.97) | 9.94<br>(7.74) | 13.18<br>(10.55) | 10.15<br>(9.09) | 3.202, p=0.077 | p=0.035 | p=0.220 |
| **IL-1β** | 10.40<br>(6.17) | 8.69<br>(4.37) | 8.43<br>(5.33) | 7.56<br>(4.71) | 0.998, p=0.320 | p=0.091 | p=0.436 |
| **IL-18** | 109.00<br>(66.75) | 77.65<br>(49.07) | 63.45<br>(52.83) | 44.34<br>(34.88) | 2.623, p=0.108 | p=0.005 | p=0.033 |
| **Eotaxin** | 34.71<br>(17.37) | 28.59<br>(22.53) | 77.23<br>(40.49) | 64.94<br>(41.34) | .348, p=0.556 | p=0.170 | p=0.190 |
| **Basic FGF** | 90.29<br>(59.41) | 89.41<br>(57.83) | 65.98<br>(45.41) | 84.77<br>(59.49) | 3.39, p=0.068 | p=0.942 | p=0.149 |
| **VEGF** | 730.31<br>(347.70) | 637.54<br>(404.34) | 315.50<br>(380.33) | 600.82<br>(373.99) | 0.47, p=0.497 | p=0.454 | p=0.165 |
| **β-NGF** | 5.44<br>(4.13) | 3.77<br>(3.03) | 4.89<br>(3.44) | 3.38<br>(2.86) | 6.426, p=0.013 | p=0.025 | p=0.067 |
| **PDGF-BB** | 1090.72<br>(1301.76) | 906.55<br>(1127.92) | 578.86<br>(561.57) | 950.82<br>(1488.20) | 1.767, p=0.186 | p=0.445 | p=0.232 |
| **IP-10** | 546.94<br>(342.87) | 460.75<br>(309.03) | 434.81<br>(254.46) | 430.42<br>(331.76) | 2.36, p=0.127 | p=0.188 | p=0.952 |
| **IL-13** | 12.11<br>(8.80) | 10.12<br>(7.24) | 11.34<br>(7.43) | 9.06<br>(7.94) | 0.237, p=0.628 | p=0.207 | p=0.214 |
| **IL-4** | 4.61<br>(2.47) | 4.52<br>(2.62) | 4.75<br>(2.56) | 4.63<br>(3.02) | 0.69, p=0.793 | p=0.868 | p=0.865 |
| **MCP-1** | 29.01<br>(23.73) | 23.49<br>(25.34) | 27.60<br>(23.66) | 23.86<br>(22.19) | 0.12, p=0.729 | p=0.286 | p=0.464 |
| **IL-8** | 14.56<br>(8.49) | 14.04<br>(9.52) | 13.11<br>(8.35) | 14.31<br>(16.47) | 0.16, p=0.690 | p=0.790 | p=0.739 |
| **MIP-1α** | 2.64<br>(1.25) | 1.98<br>(0.84) | 2.52<br>(1.31) | 1.90<br>(0.90) | 1.35, p=0.247 | p<0.001 | p=0.007 |
| **IL-10** | 26.70<br>(14.92) | 19.43<br>(9.81) | 16.38<br>(12.48) | 16.91<br>(10.47) | 1.63, p=0.204 | p=0.004 | p=0.859 |
| **G-CSF** | 76.78<br>(49.71) | 53.22<br>(31.70) | 39.57<br>(44.60) | 44.06<br>(38.25) | 2.15, p=0.145 | p=0.003 | p=0.665 |
| **GRO-a** | 2106.35<br>(1091.12) | 1532.65<br>(932.79) | 2055.57<br>(1205.08) | 1409.91<br>(987.83) | 1.828, p=0.180 | p=0.007 | p=0.018 |
| **HGF** | 703.11<br>(360.21) | 529.15<br>(291.57) | 405.60<br>(232.98) | 366.04<br>(240.22) | 10.27, p=0.002 | p=0.007 | p=0.466 |
| **IL-1α** | 47.04<br>(28.98) | 37.45<br>(22.54) | 34.27<br>(24.81) | 31.29<br>(22.72) | 4.77, p=0.031 | p=0.056 | p=0.572 |
| **IL-3** | 1.58<br>(1.41) | 1.49<br>(1.17) | 1.56<br>(1.26) | 1.27<br>(1.39) | 0.315, p=0.576 | p=0.748 | p=0.390 |
| SCF | 112.37<br>(43.25) | 91.90<br>(37.15) | 102.04<br>(51.97) | 100.76<br>(64.26) | 3.22, p=0.075 | p=0.010 | p=0.928 |
| TRAIL | 70.36<br>(37.76) | 55.35<br>(31.87) | 81.40<br>(44.60) | 59.72<br>(31.86) | .000, p=0.984 | p=0.030 | p=0.006 |
| M-CSF | 23.96<br>(18.18) | 15.58<br>(11.35) | 17.996<br>(9.79) | 10.16<br>(7.96) | 12.63, p=<.001 | p=0.003 | p<.001 |
| CTACK | 806.00<br>(368.90) | 683.44<br>(439.07) | 704.02<br>(439.95) | 677.06<br>(475.66) | 0.78, p=0.378 | p=0.164 | p=0.800 |
| IL-15 | 1308.41<br>(608.60) | 1106.61<br>(473.62) | 1184.33<br>(457.93) | 1179.91<br>(533.16) | 4.39, p=0.042 | p=0.145 | p=0.979 |
| IL-7 | 132.06<br>(106.15) | 88.09<br>(62.09) | 146.80<br>(111.46) | 94.51<br>(63.59) | 0.95, p=0.758 | p=0.004 | p=0.003 |
| IL-12<br>(p70) | 9.72<br>(8.71) | 10.23<br>(19.63) | 6.75<br>(6.49) | 8.23<br>(9.96) | 0.785, p=0.378 | p=0.821 | p=0.548 |
| IL-17 | 35.40<br>(43.76) | 29.54<br>(41.74) | 24.50<br>(42.45) | 26.35<br>(46.14) | 0.002, p=0.967 | p=0.503 | p=0.859 |
| IL-9 | 1132.29<br>(279.01) | 1029.48<br>(268.83) | 982.46<br>(277.63) | 982.21<br>(263.99) | 3.44, p=0.066 | p=0.068 | p=0.997 |
| SCGF-β | 101902.99<br>(85150.18) | 86899.98<br>(73832.28) | 54945.93<br>(37467.66) | 51673.66<br>(41618.35) | 0.019, p=0.889 | p=0.342 | p=0.726 |
| Tryptophan<br>(TRP) $\mu\text{g/mL}$ | 10.56<br>(0.95) | 10.497<br>(0.87) | 10.70<br>(0.88) | 10.63<br>(0.94) | 0.03, p=0.862 | p=0.780 | p=0.752 |
| Kynurenine<br>(Kyn) mg/mL | 0.36<br>(0.61) | 0.36<br>(0.06) | 0.35<br>(0.06) | 0.36<br>(0.42) | 0.001, p=0.973 | p=0.805 | p=0.567 |
| 3-hydroxykynurenine<br>(3-HK) ng/mL | 42.56<br>(4.78) | 41.63<br>(5.54) | 40.95<br>(5.45) | 42.04<br>(6.19) | 0.007, p=0.935 | p=0.413 | p=0.445 |
| Quinolinic acid<br>(QUIN) ng/mL | 78.76<br>(8.53) | 77.09<br>(6.09) | 77.92<br>(5.80) | 78.07<br>(5.35) | 0.51, p=0.475 | p=0.367 | p=0.908 |
| Kynurenic acid<br>(KYNA) ng/mL | 8.15<br>(1.33) | 8.61<br>(1.24) | 8.90<br>(1.28) | 8.57<br>(1.21) | 0.04, p=0.841 | p=0.133 | p=0.259 |
| Kyn/TRP ratio *1000 | 34.03<br>(7.32) | 34.36<br>(6.35) | 33.08<br>(6.74) | 34.08<br>(4.70) | 0.002, p=0.969 | p=0.840 | p=0.617 |
| KYNA/Kyn ratio | 23.97<br>(7.18) | 25.197<br>(8.42) | 26.07<br>(12.98) | 25.12<br>(5.04) | 0.449, p=0.504 | p=0.471 | p=0.732 |
| 3-HK/Kyn ratio | 125.19<br>(35.34) | 121.83<br>(39.00) | 126.07<br>(48.96) | 116.06<br>(25.17) | 0.136, p=0.713 | p=0.684 | p=0.352 |
| KYNA/QUIN ratio | 0.10<br>(0.11) | 0.11<br>(0.02) | 0.11<br>(0.12) | 0.11<br>(0.19) | 0.09, p=0.764 | p=0.101 | p=0.111 |

**Supplementary Table 3.** Signaling molecules circulating levels at the third trimester significantly associated with antepartum depression at PLS regression, and at single GLZM models.

| Rank | Variable | PLS regression |  | GLZM |  |
| --- | --- | --- | --- | --- | --- |
| | | VIP | w | LR$\chi^2$ | p |
| 1 | MIP1α (CCL3) | 1.871 | 0.289 | 8.485 | 0.0036 |
| 2 | HGF | 1.737 | 0.268 | 7.419 | 0.0064 |
| 3 | β-NGF | 1.639 | 0.253 | 5.440 | 0.0197 |
| 4 | IL-13 | 1.628 | 0.251 | 7.139 | 0.0075 |
| 5 | LIF | 1.498 | 0.231 | 5.634 | 0.0176 |
| 6 | IL-2 | 1.442 | 0.222 | 4.698 | 0.0302 |
| 7 | M-CSF | 1.241 | 0.192 | 3.311 | 0.0688 |
| 8 | IL-12 | 1.178 | 0.182 | 3.313 | 0.0687 |
| 9 | IL-3 | 1.165 | 0.180 | 3.436 | 0.0638 |
| 10 | SDF1 $\alpha$ | 1.165 | 0.180 | 3.633 | 0.0566 |
| 11 | MCP-1 | 1.156 | 0.178 | 3.168 | 0.0751 |
| 12 | MCP-3 | 1.141 | 0.176 | 3.208 | 0.0733 |
| 13 | IL1ra | 1.136 | 0.175 | 3.229 | 0.0723 |
| 14 | GRO $\alpha$ (CXCL1) | 1.132 | 0.175 | 2.990 | 0.0838 |
| 15 | IL1α | 1.097 | 0.169 | 3.027 | 0.0819 |
| 16 | IL12 p70 | 1.089 | 0.168 | 2.878 | 0.0898 |
| 17 | TRAIL | 1.078 | 0.166 | 2.923 | 0.0873 |
| 18 | TNF-β (LT- $\alpha$) | 1.033 | 0.159 | 2.724 | 0.0989 |
| 19 | IL-8 | 1.015 | 0.157 | 2.659 | 0.1030 |

## Notes

### Competing Interest Statement

The authors have declared no competing interest.

### Author Declarations

The Comitato Etico Territoriale Lombardia 1 of the IRCCS Ospedale San Raffaele and the Comitato Etico Territoriale Lombardia 3 of the Fondazione IRCCS Ca' Granda Ospedale Maggiore Policlinico gave ethical approval for this work.

## REFERENCES

1. Al-Abri K, Edge D, Armitage CJ (2023): Prevalence and correlates of perinatal depression. Soc Psychiatry Psychiatr Epidemiol. 58:1581–1590.

2. Ferrari AJ, Miller PA, Shadid J, Herrera AM, Ahmed SM, Malpani P, et al. (2026): The global prevalence of major depressive disorder during the peripartum period: a systematic review and meta-regression. The Lancet Psychiatry.

3. Chen J-H, Luo Q-M, Tu Y-Y (2026): Real world pharmacovigilance assessment of drug related perinatal depression risks. Frontiers in Pharmacology. 17:1820606.

4. Chin K, Wendt A, Bennett IM, Bhat A (2022): Suicide and maternal mortality. Curr Psychiatry Rep. 24:239–275.

5. Paulson JF, Dauber S, Leiferman JA (2006): Individual and combined effects of postpartum depression in mothers and fathers on parenting behavior. Pediatrics. 118:659–668.

6. Redic MM, Luby JL, Constantino-Pettit AM, Triplett RL, Gerstein ED, Ruscitti M, et al. (2025): Examining Associations Between the Timing of Maternal Depressive Symptoms, Parenting Behaviors, and Child Outcomes. J Am Acad Child Adolesc Psychiatry.

7. Rogers A, Obst S, Teague SJ, Rossen L, Spry EA, Macdonald JA, et al. (2020): Association between maternal perinatal depression and anxiety and child and adolescent development: a meta-analysis. JAMA pediatrics. 174:1082–1092.

8. Morais A, Pasion R, Pinto TM, Ciuffo G, Ionio C, Costa R, et al. (2025): Perinatal anxiety and depressive symptoms and maternal parenting behavior during the first three years postpartum: a systematic review. Depress Anxiety. 2025:1801371.

9. Drexhage HA, Bergink V, Poletti S, Benedetti F, Osborne LM (2025): Conventional and new immunotherapies for immune system dysregulation in postpartum mood disorders: comparisons to immune system dysregulations in bipolar disorder, major depression, and postpartum autoimmune thyroid disease. Expert Rev Clin Immunol. 21:113–135.

10. Sawyer KM (2021): The role of inflammation in the pathogenesis of perinatal depression and offspring outcomes. Brain, behavior, & immunity-health. 18:100390.

11. Okagbue HI, Adamu PI, Bishop SA, Oguntunde PE, Opanuga AA, Akhmetshin EM (2019): Systematic review of prevalence of antepartum depression during the trimesters of pregnancy. Open access Macedonian journal of medical sciences. 7:1555.

12. Silva-Fernandes A, Conde A, Marques M, Caparros-Gonzalez RA, Fransson E, Mesquita AR, et al. (2024): Inflammatory biomarkers and perinatal depression: A systematic review. PLoS ONE. 19:e0280612.

13. Sha Q, Madaj Z, Keaton S, Escobar Galvis ML, Smart L, Krzyzanowski S, et al. (2022): Cytokines and tryptophan metabolites can predict depressive symptoms in pregnancy. Translational psychiatry. 12:35.

14. Mancuso RA, Ross KM, Accortt E, Coussons-Read M, Okun ML, Irwin J, et al. (2024): Prenatal mood and anxiety disorders and associated cytokine changes. Journal of affective disorders. 347:635–644.

15. Maes M, Abe Y, Sirichokchatchawan W, Suwimonteerabutr J, Sangkomkamhangd U, Almulla AF, et al. (2023): The cytokine, chemokine, and growth factor network of prenatal depression. Brain sciences. 13:727.

16. Karlsson L, Nousiainen N, Scheinin NM, Maksimow M, Salmi M, Lehto SM, et al. (2017): Cytokine profile and maternal depression and anxiety symptoms in mid-pregnancy—the FinnBrain Birth Cohort Study: L. Karlsson et al. Arch Womens Ment Health. 20:39–48.

17. Leff Gelman P, Mancilla-Herrera I, Flores-Ramos M, Saravia Takashima M, Cruz Coronel F, Cruz Fuentes C, et al. (2019): The cytokine profile of women with severe anxiety and depression during pregnancy. BMC Psychiatry. 19:104.

18. Achtyes E, Keaton SA, Smart L, Burmeister AR, Heilman PL, Krzyzanowski S, et al. (2020): Inflammation and kynurenine pathway dysregulation in post-partum women with severe and suicidal depression. Brain, behavior, and immunity. 83:239–247.

19. Liu H, Zhang Y, Gao Y, Zhang Z (2016): Elevated levels of Hs-CRP and IL-6 after delivery are associated with depression during the 6 months post partum. Psychiatry Res. 243:43–48.

20. Corwin EJ, Pajer K, Paul S, Lowe N, Weber M, McCarthy DO (2015): Bidirectional psychoneuroimmune interactions in the early postpartum period influence risk of postpartum depression. Brain, behavior, and immunity. 49:86–93.

21. Ono CT, Yu Z, Obara T, Ishikuro M, Murakami K, Kikuya M, et al. (2023): Association between low levels of anti-inflammatory cytokines during pregnancy and postpartum depression. Psychiatry Clin Neurosci. 77:434–441.

22. Simpson W, Steiner M, Coote M, Frey BN (2016): Relationship between inflammatory biomarkers and depressive symptoms during late pregnancy and the early postpartum period: a longitudinal study. Brazilian Journal of Psychiatry. 38:190–196.

23. Nazzari S, Molteni M, Valtorta F, Comai S, Frigerio A (2020): Prenatal IL-6 levels and activation of the tryptophan to kynurenine pathway are associated with depressive but not anxiety symptoms across the perinatal and the post-partum period in a low-risk sample. Brain, Behavior, and Immunity. 89:175–183.

24. Osborne LM, Monk C (2013): Perinatal depression—the fourth inflammatory morbidity of pregnancy?: theory and literature review. Psychoneuroendocrinology. 38:1929–1952.

25. Modzelewski S, Oracz A, Iłendo K, Sokół A, Waszkiewicz N (2023): Biomarkers of postpartum depression: a narrative review. Journal of clinical medicine. 12:6519.

26. Hodges TE, Brown HK, Bodnar TS, Galea LA (2025): Inflammatory signalling during the perinatal period: implications for short-and long-term disease risk. Psychoneuroendocrinology. 172:107245.

27. Savitz J, Drevets WC, Wurfel BE, Ford BN, Bellgowan PS, Victor TA, et al. (2015): Reduction of kynurenic acid to quinolinic acid ratio in both the depressed and remitted phases of major depressive disorder. Brain, behavior, and immunity. 46:55–59.

28. Hoekzema E, Barba-Müller E, Pozzobon C, Picado M, Lucco F, García-García D, et al. (2017): Pregnancy leads to long-lasting changes in human brain structure. Nat Neurosci. 20:287–296.

29. Lisofsky N, Gallinat J, Lindenberger U, Kühn S (2019): Postpartal Neural Plasticity of the Maternal Brain: Early Renormalization of Pregnancy-Related Decreases? Neurosignals. 27:12–24.

30. Luders E, Gaser C, Gingnell M, Engman J, Sundström Poromaa I, Kurth F (2021): Gray matter increases within subregions of the hippocampal complex after pregnancy. Brain Imaging Behav. 15:2790–2794.

31. Oatridge A, Holdcroft A, Saeed N, Hajnal JV, Puri BK, Fusi L, et al. (2002): Change in brain size during and after pregnancy: study in healthy women and women with preeclampsia. AJNR Am J Neuroradiol. 23:19–26.

32. Pritschet L, Taylor CM, Cossio D, Faskowitz J, Santander T, Handwerker DA, et al. (2024): Neuroanatomical changes observed over the course of a human pregnancy. Nat Neurosci. 27:2253–2260.

33. Barba-Müller E, Craddock S, Carmona S, Hoekzema E (2019): Brain plasticity inpregnancy and the postpartum period: links to maternal caregiving and mental health. Arch Womens Ment Health. 22:289–299.

34. Kim P, Leckman JF, Mayes LC, Feldman R, Wang X, Swain JE (2010): The plasticity of human maternal brain: longitudinal changes in brain anatomy during the early postpartum period. Behav Neurosci. 124:695–700.

35. Li Y, Chu T, Che K, Dong F, Shi Y, Ma H, et al. (2021): Abnormalities of cortical structures in patients with postpartum depression: A surface-based morphometry study. Behav Brain Res.410:113340.

36. Chen C, Li B, Zhang S, Liu Z, Wang Y, Xu M, et al. (2023): Aberrant structural and functional alterations in postpartum depression: a combined voxel-based morphometry and resting-state functional connectivity study. Front Neurosci. 17:1138561.

37. Cheng B, Hu X, Roberts N, Zhao Y, Xu X, Zhou Y, et al. (2022): Prolactin mediates the relationship between regional gray matter volume and postpartum depression symptoms. J Affect Disord. 301:253–259.

38. Wise T, Radua J, Via E, Cardoner N, Abe O, Adams TM, et al. (2017): Common and distinct patterns of grey-matter volume alteration in major depression and bipolar disorder: evidence from voxel-based meta-analysis. Mol Psychiatry. 22:1455–1463.

39. Marsland AL, Gianaros PJ, Abramowitch SM, Manuck SB, Hariri AR (2008): Interleukin-6 covaries inversely with hippocampal grey matter volume in middle-aged adults. Biol Psychiatry. 64:484–490.

40. Cena L, Mirabella F, Palumbo G, Gigantesco A, Trainini A, Stefana A (2021): Prevalence of maternal antenatal and postnatal depression and their association with sociodemographic and socioeconomic factors: A multicentre study in Italy. J Affect Disord. 279:217–221.

41. Stefana A, Cena L, Alice T, Palumbo G, Gigantesco A, Mirabella F (2024): The diagnostic accuracy of the Edinburgh Postnatal Depression Scale without the self-harm item: Does culture matter?, Journal of Psychiatric Research. J Psychiatr Res. 175:432–434.

42. Comai S, Melloni E, Lorenzi C, Bollettini I, Vai B, Zanardi R, et al. (2022): Selective association of cytokine levels and kynurenine/tryptophan ratio with alterations in white matter microstructure in bipolar but not in unipolar depression. Eur Neuropsychopharmacol. 55:96–109.

43. Simonato M, Dall’Acqua S, Zilli C, Sut S, Tenconi R, Gallo N, et al. (2021): Tryptophan Metabolites, Cytokines, and Fatty Acid Binding Protein 2 in Myalgic Encephalomyelitis/Chronic Fatigue Syndrome. Biomedicines. 9.

44. Liu C, Zhang X, Nguyen TT, Liu J, Wu T, Lee E, et al. (2022): Partial least squares regression and principal component analysis: similarity and differences between two popular variable reduction approaches. Gen Psychiatr. 35:e100662.

45. Akarachantachote N, Chadcham S, Saithanu K (2014): Cutoff threshold of variable importance in projection for variable selection. Int J Pure Appl Math. 94:307–322.

46. Chong I-G, Jun C-H (2005): Performance of some variable selection methods when multicollinearity is present. Chemometrics and intelligent laboratory systems. 78:103–112.

47. Hill T, Lewicki P, Lewicki P (2006): Statistics: methods and applications: a comprehensive reference for science, industry, and data mining. StatSoft, Inc.

48. Palermo G, Piraino P, Zucht H-D (2009): Performance of PLS regression coefficients in selecting variables for each response of a multivariate PLS for omics-type data. Advances and Applications in Bioinformatics and Chemistry.57–70.

49. McCullagh P, Nelder JA (1989): Generalized linear models (2nd Ed.). New York: Chapman & Hall.

50. Agresti A (1996): An Introduction to Categorical Data Analysis. New York: Wiley.

51. Dobson AJ (1990): An introduction to generalized linear models. New York: Chapman & Hall.

52. Poletti S, Zanardi R, Mandelli A, Aggio V, Finardi A, Lorenzi C, et al. (2024): Low-dose interleukin 2 antidepressant potentiation in unipolar and bipolar depression: Safety, efficacy, and immunological biomarkers. Brain Behav Immun. 118:52–68.

53. Bravi B, Verga C, Palladini M, Poletti S, Buticchi C, Stefania S, et al. (2025): Effects of kynurenine pathway metabolites on choroid plexus volume, hemodynamic response, and spontaneous neural activity: A new mechanism for disrupted neurovascular communication and impaired cognition in mood disorders. Brain Behav Immun. 125:414–427.

54. Harrington YA, Fortaner-Uyà L, Paolini M, Poletti S, Lorenzi C, Spadini S, et al. (2024): Disentangling the Genetic Landscape of Peripartum Depression: A Multi-Polygenic Machine Learning Approach on an Italian Sample. Genes (Basel). 15.

55. Harrington YA, Paolini M, Fortaner-Uyà L, Maccario M, Melloni EMT, Poletti S, et al. (2025): History of Peripartum Depression Moderates the Association Between Estradiol Polygenic Risk Scores and Basal Ganglia Volumes in Major Depressive Disorder. Biol Psychiatry Cogn Neurosci Neuroimaging. 10:7–16.

56. Breiman L, Friedman J, Olshen RA, Stone CJ (2017): Classification and regression trees. Chapman and Hall/CRC.

57. Hill T, Lewicki P (2006): Statistics: methods and applications. A comprehensive reference for science, industry, and data mining. General Linear Models, StatSoft, Tulsa (OK). Chapter 18:245–276.

58. PrabhuDas M, Bonney E, Caron K, Dey S, Erlebacher A, Fazleabas A, et al. (2015): Immune mechanisms at the maternal-fetal interface: perspectives and challenges. Nat Immunol. 16:328–334.

59. Ysrraelit MC, Correale J (2019): Impact of sex hormones on immune function and multiple sclerosis development. Immunology. 156:9–22.

60. Murgia F, Giagnoni F, Lorefice L, Caria P, Dettori T, D’Alterio MN, et al. (2022): Sex hormones as key modulators of the immune response in multiple sclerosis: a review. Biomedicines. 10:3107.

61. Sha Q, Galvis MLE, Madaj ZB, Keaton SA, Smart L, Edgerly YM, et al. (2024): Dysregulated placental expression of kynurenine pathway enzymes is associated with inflammation and depression in pregnancy. Brain, behavior, and immunity. 119:146–153.

62. Poletti S, Mazza MG, Benedetti F (2024): Inflammatory mediators in major depression and bipolar disorder. Translational Psychiatry. 14:247.

63. Poletti S, Melloni E, Aggio V, Colombo C, Valtorta F, Benedetti F, et al. (2019): Grey and white matter structure associates with the activation of the tryptophan to kynurenine pathway in bipolar disorder. J Affect Disord. 259:404–412.

64. Poletti S, Vai B, Mazza MG, Zanardi R, Lorenzi C, Calesella F, et al. (2021): A peripheral inflammatory signature discriminates bipolar from unipolar depression: a machine learning approach. Prog Neuropsychopharmacol Biol Psychiatry. 105:110136.

65. Han KM, Ham BJ (2021): How Inflammation Affects the Brain in Depression: A Review of Functional and Structural MRI Studies. J Clin Neurol. 17:503–515.

66. Bai YM, Chen MH, Hsu JW, Huang KL, Tu PC, Chang WC, et al. (2020): A comparison study of metabolic profiles, immunity, and brain gray matter volumes between patients with bipolar disorder and depressive disorder. J Neuroinflammation. 17:42.

67. Chen MH, Chang WC, Hsu JW, Huang KL, Tu PC, Su TP, et al. (2019): Correlation of proinflammatory cytokines levels and reduced gray matter volumes between patients with bipolar disorder and unipolar depression. J Affect Disord. 245:8–15.

68. Fusar-Poli P, Placentino A, Carletti F, Landi P, Allen P, Surguladze S, et al. (2009): Functional atlas of emotional faces processing: a voxel-based meta-analysis of 105 functional magnetic resonance imaging studies. J Psychiatry Neurosci. 34:418–432.

69. Li Y, Chu T, Che K, Dong F, Shi Y, Ma H, et al. (2021): Altered gray matter structural covariance networks in postpartum depression: a graph theoretical analysis. J Affect Disord. 293:159–167.

70. Salgado G, Chaves C, Salgado A, Palacios B, Duque A (2026): Attentional biases in women with postpartum depressive symptoms: A comparative eye-tracking study. J Affect Disord. 401:121263.

71. Schwarcz R, Bruno JP, Muchowski PJ, Wu HQ (2012): Kynurenines in the mammalian brain: when physiology meets pathology. Nat Rev Neurosci. 13:465–477.

72. Stone TW, Clanchy FIL, Huang YS, Chiang NY, Darlington LG, Williams RO (2022): An integrated cytokine and kynurenine network as the basis of neuroimmune communication. Front Neurosci. 16:1002004.

73. Barone P (2019): The ‘Yin’and the ‘Yang’of the kynurenine pathway: Excitotoxicity and neuroprotection imbalance in stress-induced disorders. Behavioural pharmacology. 30:163–186.

74. Mastria G, Scaliti E, Mehring C, Burdet E, Becchio C, Serino A, et al. (2023): Morphology, Connectivity, and Encoding Features of Tactile and Motor Representations of the Fingers in the Human Precentral and Postcentral Gyrus. J Neurosci. 43:1572–1589.

75. Drake PM, Gunn MD, Charo IF, Tsou CL, Zhou Y, Huang L, et al. (2001): Human placental cytotrophoblasts attract monocytes and CD56(bright) natural killer cells via the actions of monocyte inflammatory protein 1alpha. J Exp Med. 193:1199–1212.

76. Laham N, Van Dunné F, Abraham LJ, Farrugia W, Bendtzen K, Brennecke SP, et al. (1997): Tumor necrosis factor-beta in human pregnancy and labor. J Reprod Immunol. Ireland, pp 53–69.

77. Alijotas-Reig J, Llurba E, Gris JM (2014): Potentiating maternal immune tolerance in pregnancy: a new challenging role for regulatory T cells. Placenta. 35:241–248.

78. Zhou L (2025): The Mechanisms and therapeutic effects of granulocyte colony-stimulating factor in reproduction. Reprod Biol Endocrinol. 23:78.

79. Vondra S, Höbler AL, Lackner AI, Raffetseder J, Mihalic ZN, Vogel A, et al. (2023): The human placenta shapes the phenotype of decidual macrophages. Cell Rep. 42:111977.

80. Horii Y, Ino H, Sasaki F, Ishibashi M, Koike E, Kawamoto Y, et al. (2025): Inflammatory cytokine interleukin-18 prevents murine miscarriage by inducing appropriate inflammation. Sci Rep. 15:30765.

81. Sharma S, Godbole G, Modi D (2016): Decidual Control of Trophoblast Invasion. Am J Reprod Immunol. 75:341–350.

82. Ma C, Liu G, Liu W, Xu W, Li H, Piao S, et al. (2021): CXCL1 stimulates decidual angiogenesis via the VEGF-A pathway during the first trimester of pregnancy. Mol Cell Biochem. 476:2989–2998.

83. Kang X, Zhang X, Liu Z, Xu H, Wang T, He L, et al. (2016): CXCR2-Mediated Granulocytic Myeloid-Derived Suppressor Cells’ Functional Characterization and Their Role in Maternal Fetal Interface. DNA Cell Biol. 35:358–365.

84. Kreicberga I, Junga A, Pilmane M (2021): Assessment of apoptosis and appearance of hepatocyte growth factor in placenta at different gestational ages: A cross-sectional study. Int J Reprod Biomed. 19:505–514.

85. Hübel J, Hieronymus T (2015): HGF/Met-Signaling Contributes to Immune Regulation by Modulating Tolerogenic and Motogenic Properties of Dendritic Cells. Biomedicines. 3:138–148.

86. Thornton CA, Holloway JA, Shute JK, Holloway JW, Diaper ND, Warner JO (2009): Human mid-gestation amniotic fluid contains interleukin-16 bioactivity. Immunology. 126:543–551.

87. Guan S, Bai X, Ding J, Zhuang R (2023): Circulating inflammatory cytokines and hypertensive disorders of pregnancy: a two-sample Mendelian randomization study. Front Immunol. 14:1297929.

88. Romanowska-Próchnicka K, Felis-Giemza A, Olesińska M, Wojdasiewicz P, Paradowska-Gorycka A, Szukiewicz D (2021): The Role of TNF-α and Anti-TNF-α Agents during Preconception, Pregnancy, and Breastfeeding. Int J Mol Sci. 22.

89. Siwetz M, Blaschitz A, El-Heliebi A, Hiden U, Desoye G, Huppertz B, et al. (2016): TNF-α alters the inflammatory secretion profile of human first trimester placenta. Lab Invest. 96:428–438.

90. Ueshima C, Kataoka TR, Osakabe M, Sugimoto A, Ushirokawa A, Shibata Y, et al. (2022): Decidualization of Stromal Cells Promotes Involvement of Mast Cells in Successful Human Pregnancy by Increasing Stem Cell Factor Expression. Front Immunol. 13:779574.

91. Khodadi E, Shahrabi S, Shahjahani M, Azandeh S, Saki N (2016): Role of stem cell factor in the placental niche. Cell Tissue Res. 366:523–531.

92. D’Angelo A, Ceccanti M, Petrella C, Greco A, Tirassa P, Rosso P, et al. (2020): Role of neurotrophins in pregnancy, delivery and postpartum. Eur J Obstet Gynecol Reprod Biol. 247:32–41.

93. Kaihola H, Olivier J, Poromaa IS, Åkerud H (2015): The effect of antenatal depression and selective serotonin reuptake inhibitor treatment on nerve growth factor signaling in human placenta. PLoS One. 10:e0116459.

94. Hannan NJ, Jones RL, White CA, Salamonsen LA (2006): The chemokines, CX3CL1, CCL14, and CCL4, promote human trophoblast migration at the feto-maternal interface. Biol Reprod.74:896–904.

95. Bevilacqua E, Paulesu L, Ferro EA, Ietta F, Faria MR, Lorenzon AR, et al. (2014): Review: putative roles for the macrophage migratory inhibitory factor at the maternal fetal interface. Placenta. 35 Suppl:S51–56.

96. Jovanović Krivokuća M, Vilotić A, Stefanoska I, Bojić-Trbojević Ž, Vićovac L (2021): Macrophage migration inhibitory factor in human early pregnancy events and association with placental pathologies. Placenta. 116:51–57.

97. Piao HL, Tao Y, Zhu R, Wang SC, Tang CL, Fu Q, et al. (2012): The CXCL12/CXCR4 axis is involved in the maintenance of Th2 bias at the maternal/fetal interface in early human pregnancy. Cell Mol Immunol. 9:423–430.

98. Phillips TA, Ni J, Pan G, Ruben SM, Wei YF, Pace JL, et al. (1999): TRAIL (Apo-2L) and TRAIL receptors in human placentas: implications for immune privilege. J Immunol. 162:6053–6059.

99. Saito S, Shima T, Nakashima A, Shiozaki A, Ito M, Sasaki Y (2007): What is the role of regulatory T cells in the success of implantation and early pregnancy? J Assist Reprod Genet. 24:379–386.

100. Scott LM, Bryant AH, Rees A, Down B, Jones RH, Thornton CA (2017): Production and regulation of interleukin-1 family cytokines at the materno-fetal interface. Cytokine. 99:194–202.

101. Hamelin-Morrissette J, Dallagi A, Girouard J, Ravelojaona M, Oufqir Y, Vaillancourt C, et al. (2020): Leukemia inhibitory factor regulates the activation of inflammatory signals in macrophages and trophoblast cells. Mol Immunol. 120:32–42.

102. Equils O, Kellogg C, McGregor J, Gravett M, Neal-Perry G, Gabay C (2020): The role of the IL-1 system in pregnancy and the use of IL-1 system markers to identify women at risk for pregnancy complications†. Biol Reprod. 103:684–694.

103. Yadav AK, Chaudhari H, Warke H, Shah PK, Dodagatta-Marri E, Kishore U, et al. (2014): Differential expression of collectins in human placenta and role in inflammation during spontaneous labor. PLoS One. 9:e108815.

